# Multimodal single-cell spatial profiling reveals altered T cell immunity and B-cell follicular architecture in non-metastatic lymph nodes of advanced NSCLC patients

**DOI:** 10.64898/2026.01.12.25343268

**Authors:** Zhan Hao Xi, Yusuke Koga, Shannon McDermott, Erin E. Kane, Roxanna Pfefferkorn, Ehab Billatos, Paul R. Hosking, Jennifer E. Beane, Eric J. Burks, Sarah A. Mazzilli, Kei Suzuki, Joshua D. Campbell

## Abstract

Regional lymph nodes (LNs) in the thoracic cavity are essential immunological hubs that coordinate humoral and cell-mediated immunity during non-small cell lung cancer (NSCLC) progression. To investigate immune dysregulation in non-metastatic regional LNs from patients with advanced-stage NSCLC, we performed multimodal profiling of 36 LNs from 11 patients undergoing curative-intent resection using CITE-seq, scRNA-seq, and Imaging Mass Cytometry (IMC). Regional N1 LNs from our advanced-stage patients (stage IB–IIIA) displayed visceral pleural invasion (VPI) or lymphovascular invasion (LVI), and exhibited significant enrichment of dysfunctional CD8⁺ T cells and regulatory T cells compared with N2 LNs and stage IA LNs. These subsets spatially co-localized with mature regulatory dendritic cells (CD1c⁺, TIM3⁺, LAMP3⁺), forming an immunosuppressive niche enriched in advance-stage N1 LNs. Concurrently, advanced-stage N1 LNs contained more “decorticated” B-cell follicles characterized by decreased encapsulation of the mantle zone layer surrounding the germinal centers. Together, these findings reveal parallel disruption of humoral and cell-mediated immunity in regional LNs of advanced-stage NSCLC patients.

**Significance:** This manuscript demonstrates that non-metastatic regional lymph nodes in advanced NSCLC are not immunologically normal but instead show coordinated remodeling of T cell immunity and B-cell follicular architecture. By integrating single-cell and spatial profiling, it identifies dysfunctional CD8 T cells, regulatory T cells, mregDC-associated suppressive niches, and disrupted follicular organization as features of early nodal immune dysregulation before overt metastasis. These findings support regional lymph nodes as clinically relevant immune sites that may improve risk stratification and guide future perioperative or adjuvant immunotherapy strategies in resected NSCLC.

## INTRODUCTION

Significant progress in the diagnosis and screening of non-small cell lung cancer (NSCLC) over the past decades has improved early detection and treatment strategies, leading to better outcomes for patients^1^. Despite these efforts, the 5-year survival rate for all stage NSCLCs remains below 30%, which is substantially lower than that of other common cancers such as breast cancer (91%) and prostate cancer (97.5%)^2^. The stage at diagnosis remains one of the most critical determinants of survival. The five-year survival rate in the United States for localized (stage I-II) disease is estimated to be 70%, whereas the survival for distant (stage IV) disease is estimated to be 12%^2–4^. This stark contrast underscores the urgent need to understand the components driving tumor aggressiveness and metastasis. Lymph node (LN) metastasis is an important prognostic indicator in the staging of NSCLC, guiding both survival estimates and treatment strategies^5^. However, pathologic assessments of the LN are limited to presence or absence of tumor cells in the N1 (hilar LNs), N2 (mediastinal LNs), and N3 (contralateral) nodal stations for therapeutic decision-making^6^. Despite the common acquisition of LN tissue during clinical workflows, aberrant immune alterations in the LNs have not been regularly used in the clinic to aid in the diagnosis and staging of NSCLC.

LNs are integral to the immune response, serving as hubs where innate immunity trains the adaptive immunity, consisting of humoral immunity (B cells) and cell-mediated immunity (CD8^+^ T cells), to respond against pathogens and malignant cells while maintaining self-tolerance^7,8^. LNs are architecturally compartmentalized into cortical (B cell follicular), paracortical (T cell), and medullary zones containing antigen-presenting cells (APCs), each supporting distinct immune functions and cellular interactions^9^. APCs such as dendritic cells (DCs) and macrophages constantly traffic to and within the LN, enabling efficient antigen capture, presentation, and subsequent T and B cell priming^10^. Helper T cells assist B cells in generating high-affinity antibodies within germinal centers, mounting effective humoral response^11–13^. Simultaneously, naïve CD8^+^ T cells can mature into effector cytotoxic T cells, be trafficked to inflammatory sites via cytokine and chemokine signaling, and target malignant cells displaying the cognate antigen^14,15^. However, aberrations in these processes can ultimately tip the balance from tumor elimination to immune tolerance and cancer progression^16–19^.

To avoid immunosurveillance, tumors can promote immune dysfunction by a variety of mechanisms including increased infiltration of immunosuppressive regulatory T cells (Tregs) or activation of inhibitory pathways on CD8^+^ T cells via up-regulation of checkpoint proteins such as LAG3, CTLA4, CD39, PD-1, TIM3, and TIGIT^20,21^. This phenotype correlates with diminished immune effector functions, production of immunosuppressive environment, and worse patient outcomes^18,22–24^. Clinical studies of pre-metastatic LNs in breast and colorectal cancer patients have identified immunosuppressive features, including the accumulation of Tregs, CD163^+^ macrophages, dysfunctional dendritic cells, and exhausted CD8^+^ T cells^20,21^. In NSCLC, deep phenotyping of the immune microenvironment has been performed in the tumor tissue to identify immunosuppressive and exhausted cell types, such as terminally exhausted CD8^+^ T cells, Tregs, SPP1^+^ immunosuppressive macrophages, LAMP3^+^ mregDCs, and B cell structures such as mature Tertiary Lymphoid Structures (TLS)^22,25–35^. However, extensive characterization of the immune dysregulation that occurs in the tumor-adjacent LNs has not been performed.

To decipher the immune alterations in the regional, non-metastatic LNs that occur in advanced stage NSCLC, we deeply characterized the immunological changes that occur between treatment-naïve patients with lower- and higher-stage disease. Single cell RNA sequencing (scRNA-seq) with Cellular Indexing of Transcriptomes and Epitopes sequencing (CITE-seq) were employed to simultaneously quantify the expression of RNA and cell-surface proteins using barcoded antibody-derived tags (ADTs) on single cells^36^. Additionally, profiling with imaging mass cytometry (IMC) was used to spatially identify aberrant immune neighborhood niches within the disrupted LN architecture^15,37^. Multimodal profiling revealed increased immunosuppressive cell populations within N1 LN regions in higher-stage patients (stage IB-IIIA) that have tumor invasive features of either Visceral Pleural Invasion (VPI) or Lymphovascular Invasion (LVI), compared to N2 LNs or LNs from individuals with lower-stages (stage IA) without tumor invasive features (**Supplementary Table 1**). Overall, these results suggest that alterations in the humoral and cell-mediated immunity pathways within the regional LNs define a permissive microenvironment enabling advanced stage NSCLC.

## RESULTS

### Characterization of the LN immune landscape in NSCLC using multimodal single-cell profiling

Tissue specimens were collected following resection from 11 patients undergoing curative-intent surgery for confirmed or suspected early-stage NSCLC (**Fig. 1A, Supplementary Tables 1, 2**). The patient cohort was dichotomized based on pathological stage and tumor invasive features, into “higher” (pStage ≥ IB) and “lower” (pStage IA) severity groups by the 8^th^ edition of the TNM staging. Fine needle aspirate (FNA) samples were obtained from one to four mediastinal (N2), hilar, and/or interlobar LNs (N1) from each patient in addition to tumor tissue for scRNA-seq and CITE-seq (**Fig. 1B**). Adjacent non-malignant tissue was also obtained for 4 patients. Samples were multiplexed using hashtag ADTs for samples from the same individual. In addition to the hashtags, we also incubated each pooled sample with the TotalSeq-C CITE-seq panel consisting of 128 antibodies targeting cell surface proteins important for characterization of immune cell populations and activity (**Methods, Supplementary Table 3**). Typically, droplets from single-cell microfluidic systems are considered “empty” or contain a true cell based on the total number of RNA counts. We recovered a subpopulation of cells that had high levels of total ADT counts but low levels of total RNA counts (**Supplementary Fig. 1A**). This population was distinct from the spongelets, a recently described artifact in CITE-seq data and instead showed specific protein level of CD16, suggesting a neutrophil identity^38^ (**Supplementary Fig. 1B**). After filtering for percentage of mitochondrial gene counts, ambient RNA contamination, and minimum expression of CITE-seq features, we analyzed a total of 85,869 single cells including 44,420 cells from the tumor, 4,857 cells from non-metastatic N1 LNs, 317 cells from metastatic N1 LNs, 30,417 cells from non-metastatic N2 LNs, 5,858 cells from the normal adjacent lung.

Unsupervised clustering of the scRNA-seq data revealed 8 clusters of broad cell types (**Fig. 1C**) which were annotated using canonical RNA and protein markers (**Fig. 1D, Supplementary Tables 4-6**). T and B cells were more prevalent in LN samples, while myeloid cells were more frequent in tumor samples (FDR q-value < 0.05; **Fig. 1E, Supplementary Fig. 2**). Epithelial cells were specific to tumor and adjacent normal samples confirming a lack of metastasis in the profiled LNs. Cell type proportions were less variable across LN samples compared to the tumor and normal lung tissue samples (**Supplementary Fig. 3**). The average correlation between each of the 122 protein markers from the CITE-seq panel and their corresponding mRNA expression was 0.54 with a range from -0.38 to 0.95 (**Fig. 1F, Supplementary Fig. 4, Supplementary Table 7**). In general, markers for broader cell types exhibited high correlation values including CD3 for T cells, CD19 and CD20 (MS4A1) for B cells, and CD14 for myeloid cells (R > 0.7). In contrast, markers of immune cell activation and T cell exhaustion, such as CD83, CTLA4, and TIGIT, exhibited low correlation values (R < 0.2).

Fixed tissue sections for one to four non-metastatic LNs were also obtained after pathological assessment for spatial profiling with IMC (**Fig. 1B**). A panel of 40 metal-labeled antibodies was designed based on the cell types and states identified through transcriptomic and proteomic expression profiles from CITE-seq data (**Supplementary Table 3**). This panel specifically targeted broad cell types (e.g. CD68 and CD163 for macrophages, CD117 for mast cells, CD19 for B cells.), exhaustion markers (e.g. LAG3, TIGIT, TIM3), and functional or activation states (e.g. Ki-67, CD28, CD86, CD45RA, CD45RO, HLA-DRB, ITGAE). To aid in selection of regions of interest (ROIs) for IMC ablation, we first performed hematoxylin and eosin (H&E) staining, followed by multiplex immunofluorescence (mIF) using CD3E for T cells, CD19 for B cells, CD68 for macrophages, CD56 for NK cells, and CD11c for DCs (**Fig. 1G**). The mIF and H&E images were reviewed by a pathologist to assess overall cell type distribution and tissue architecture for IMC ROI ablation selection, ensuring inclusion of B cell follicles and surrounding T cell zones (**Fig. 1G**). In total, 104 ROIs across 30 LNs were profiled, with an average ablation area of ∼7mm^2^ per ROI, and a total of 265 B cell follicular regions were identified. Following IMC acquisition and cell segmentation, a total of 1,128,788 cells were obtained, clustered, and annotated into eight broad cell populations based on the expression of established protein markers, including tumor-associated macrophages (TAMs) (**Fig. 1H-I**). Consistent with CITE-seq findings, all 8 IMC cell types were detected in the LNs across all patients (**Fig. 1J, Supplementary Fig. 5**).

**Fig. 1.**
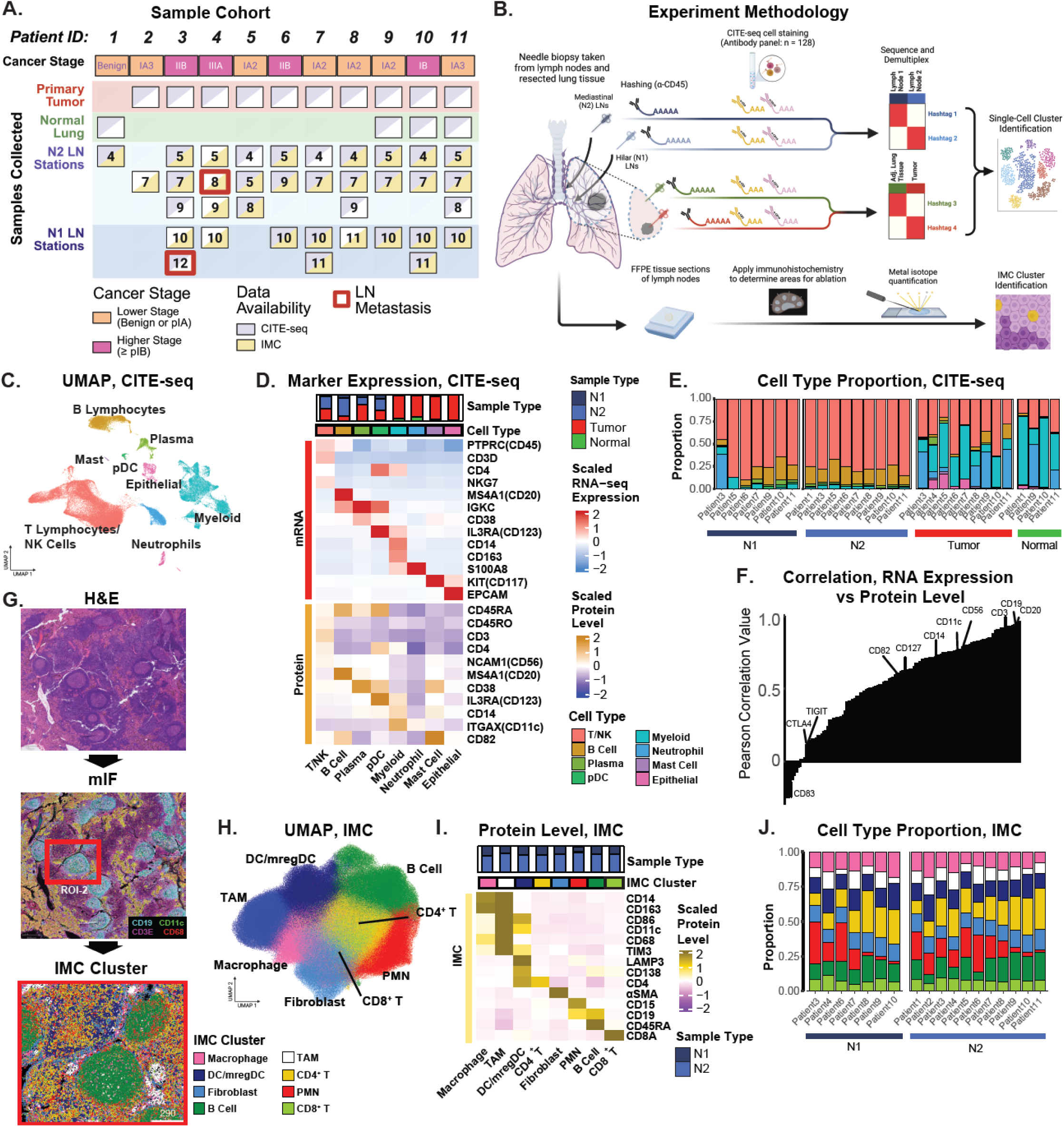
Comparing protein and RNA expression from tumor, normal, and lymph node tissue obtained from lung resections. **A)** Primary tumor, normal lung, and LN tissues were obtained from 11 individuals. Primary tumor (red) samples were collected from 10 patients with detected malignant tumors. Normal lung tissue (green) was collected from 4 patients. Additionally, a benign lesion sample was obtained from one patient with hamartoma diagnosis. Patients were divided into 3 categories based on pathological stage: benign, lower (pStage < IB), and higher (pStage ≥ IB) stage. **B)** Experimental approach (top). FNA samples were subject to scRNA-seq with CITE-seq to multiplex and label cell surface markers of immune populations. Upon sequencing, demultiplexing was conducted based on the expression of the hashtag for each cell. Demultiplexed cells are selected for downstream analysis including clustering, annotation, and identification of tissue enrichment. (bottom) IMC analysis utilized formalin-fixed paraffin-embedded (FFPE) tissue sections from the resected LN tissues, where regions of interest were selected using a 6-marker IF panel. Validation of cell type enrichment and proximity-based inference of cell-cell neighborhoods was performed on clusters of single cells identified through segmentation. **C)** UMAP projections based on RNA expression were computed across all cells. Points are annotated by broad cell type annotations (left) and originating tissue type (right). **D)** Expression of selected RNA (top) and protein (bottom) markers are shown for broad cell types. Marker expression is z-score normalized across cells for visualization. **E)** The relative proportion of cell populations per patient is given. Broadly, T and B cells were more prevalent in LN samples, while myeloid cells, neutrophils, and epithelial cells were more prevalent in tumor and normal lung tissue samples. **F)** The Pearson correlation coefficient was calculated to determine the correlation between the average RNA expression and matching protein levels for all cell populations across 122 markers in the CITE-seq panel. Markers associated with broader cell types such as CD3, CD19 and CD20 (MS4A1) were amongst the highest correlated. **G)** Representative hematoxylin and eosin (H&E; top) and immunofluorescent staining (mIF; middle) were conducted on FFPE tissue sections of LNs prior to ablation to evaluate suitable targets regions. An example of ablated ROI is denoted (red box). Single-cell masks are colored by defined IMC cluster labels (bottom). **H)** A total of 8 clusters were identified through unsupervised clustering of single segmented cells identified in the IMC analysis. UMAP projections based on antibody intensity were computed across all single cells. **I)** Expression of selected markers for imaging data-based cluster labels is shown. **J)** The relative proportion of cell type clusters per patient.

### Integration of surface protein levels improves phenotyping of lymphocyte cell states and identifies enrichment of regulatory T cells, dysfunctional CD8^+^ T cells, and plasma cells in the N1 LNs of higher-stage patients

As critical mediators of the adaptive immune response, T cells have been targeted in immunotherapeutic strategies for the treatment of NSCLC. One notable case is the use of immune checkpoint inhibitors (ICIs), such as PD-1 (pembrolizumab, nivolumab), CTLA4 (ipilimumab), and LAG3 inhibitors (relatlimab), which prevent immune checkpoint pathways from being exploited by cancer cells^39–41^. To further characterize T cell subpopulations, 47,068 cells from T cells or natural killer (NK) lineages were subclustered using their transcriptomic profiles from the scRNA-seq data (**Fig. 2A, Supplementary Fig. 6**). The refined cell clusters included 7 CD4^+^ T cell subpopulations, 5 CD8^+^ T cell subpopulations, 3 NK cell subpopulations (*NCAM1*), and 1 γδ T cell cluster (*TRDV2*, *TRGV9*; **Fig. 2B**). When examining the correlation between the RNA expression and corresponding protein level in T cells, we observed high correlation for cell type markers such as CD4 and CD8 as well as some markers for T cell activation including CD39 (*ENTPD1*), CD103 (*ITGAE*), and CD62L (*SELL*) (R > 0.7; **Fig. 2C**; **Supplementary Fig. 7**). RNA and protein marker expression for PD-1 was moderately correlated (R = 0.675). In contrast, other immune regulatory markers such as *TIGIT, LAG3,* and *CTLA4* exhibited lower correlations (R < 0.25), suggesting that RNA expression alone may not be sufficient to capture T-cell exhaustion phenotypes.

CD4^+^ T cells are essential for tumor immunity by orchestrating and sustaining effective antitumor responses through modulating CD8^+^ T cells, promoting antibody production in B cells, modulating the tumor microenvironment, and, in some cases, directly killing tumor cells^42^. Within the data, CD4^+^ T-cell populations identified included naïve (*TCF7, CD62L,* CD45RA), central memory (*KLF2*, *S1PR1*, *ITGB1*, *CD62L*, *CD127*, CD45RO), regulatory (Tregs; *FOXP3*), effector memory (*CD127*, *CD40LG*, *CD69*, CD45RO), T follicular helper (*PD-1*, *ICOS*, *CD40LG*, *TOX2*, *BATF*, *BCL6*), dysfunctional (*CTLA4*, *TIGIT*, *TIM3*, *LAG3*, *PD-1*, *ICOS*, *TOX*, *TOX2*, CD45RO), and inflammatory CD4^+^ T cells (*CXCR4*, *CD103*, *CD127*, *IFNG*, *FOS*; **Fig. 2D; Supplementary Tables 8, 9**). 5 of the 7 identified CD4^+^ T-cell subpopulations were enriched in LNs compared to the tumor including the naïve, memory, and follicular helper (Tfh) CD4^+^ T cell subpopulations (**Supplementary Fig. 8, Supplementary Table 10**). Interestingly, while the dysfunctional Th1-like and inflammatory subpopulations were significantly enriched in tumors, they were also present in low proportions in a subset of LNs. Their gene expression was similar to the CD4^+^ Tfh cells but with additional upregulation of *IFNG*, *CXCR6*, and *TIM3* gene expression, higher CD39 CITE-seq expression, as well as downregulation of *CD27* gene expression, suggesting an immunosuppressive phenotype. Tregs were re-clustered into two distinct populations (**Fig. 2D**), each exhibiting enrichment in either the tumor or the LN. LN-enriched Tregs expressed higher levels of *CD62L*, *TCF7*, *CCR7*, and CD45RA, indicative of a more naïve or central memory phenotype, while tumor-enriched Tregs showed elevated expression of activation and dysfunctional markers, including *ICOS*, *CD39*, and *PD-1*, indicative of an immunosuppressive role in the tumor microenvironment (**Fig. 2E**).

CD8^+^ T cells are critical for tumor immunity by directly recognizing tumor associated antigens and killing cancer cells through the release of cytotoxic molecules such as perforin and granzymes, and by secreting cytokines like IFN-γ to enhance the antitumor immune response^43^. In our dataset, subpopulations of CD8^+^ T cells included naïve CD8^+^ T cells (*CD62L*, *CCR7*, *LEF1*, *CD27*, *CD127*, CD45RA), effector memory CD8^+^ T cells (*CD27*, *CD127*, *GZMK*, CD45RO), mucosal-associated invariant CD8^+^ T cells (MAITs) (*KLRB1*, *GZMK*), dysfunctional CD8+ T cells (*GZMB*, *IFNG*, *CTLA4*, *TIGIT*, *TIM3*, *PD-1*, *LAG3*, *CD103*, *CD39*), and proliferating dysfunctional CD8+ T cells (*MKI67*, *STMN1*, *CTLA4*, *TIGIT*, *TIM3*, *PD-1*, *LAG3*, CD45RO; **Fig. 2F**). Three patient-specific clusters were aggregated and labeled as a single group of dysfunctional CD8^+^ T cells based on shared expression of dysfunction-associated markers. Expectedly, naïve and effector memory CD8^+^ T cells were significantly more abundant in LN samples, and MAIT was found in both tumor and benign tissue (**Supplementary Fig. 8**). Dysfunctional and proliferating dysfunctional CD8^+^ T cells were enriched in tumor tissue (FDR q-value < 0.05) compared to the LNs. NK cells are not only vital for tumor immunity by directly eliminating cancer cells without prior sensitization and shaping adaptive immune responses but also regulate and prime immunity in the LN^44^. Upon clustering of NK cells (CD56^+^, CD16^+^), two CD56^+^ CD16^-^ and one CD16^+^ CD56^-^ NK population was identified (**Supplementary Fig. 9**). Concordant with previous findings, CD56^+^ NK cells were more abundant in LNs while CD16^+^ NK cells were more abundant in tumors^45,46^ (**Supplementary Fig. 8**). The expression of granzymes and granulysins, essential for the cytolytic activity of NK cells, differed by subset, with *GZMK* being upregulated in the CD56^+^ NK cells, *GZMB* and *GZMH* upregulated in CD16^+^ NK cells, and *GNLY* upregulated in the CCR7^+^ CD56^+^ NK subset along with the CD16^+^ NK cell population.

Lastly, 9,565 B cells and plasma cells were subclustered into 8 subpopulations (**Supplementary Figs. 10, 11; Supplementary Tables 11, 12**). Similarly to the T cells, most broad cell type markers displayed a high correlation between the RNA expression and corresponding protein levels (**Supplementary Figs. 11B, 12**). Two major subsets of B cells were identified and characterized by high levels of *CCR7* or *CD38* expression **(Supplementary Fig. 11C**). Amongst the *CCR7*^+^ subpopulations were naïve B cells (*TCL1A*, *IGHD*) and memory B cells. Additionally, a subpopulation with antigen-presentation features (*CD11C*, MHC-II, CD86) was also discernable. Two types of germinal center (GC) B cells (*CD38*^+^, *BCL6* and *ELL3)* were identified, with one displaying increased CD24 expression. Plasma cells were characterized *as CD138*^+^, *JCHAIN*^+^, *CD38*^+^ cells and were identified in both LN and tumor tissue.

Differential composition analysis was conducted to determine which immune cell populations were enriched in N1 LNs from higher-stage patients compared to N2 LNs from higher-stage patients and all LNs from lower-stage patients (**Supplementary Figs. 13, 14; Supplementary Tables 13, 14**). When considering all Tregs as a single group, they were enriched in higher-stage N1 LNs (FDR = 0.03; **Fig. 2F**). Similarly, despite being an overall lower percentage of cells, the dysfunctional CD8^+^ T cells were also enriched in the higher-stage N1 LNs (FDR = 0.019; **Fig. 2F**). Within the NK cells, the CD56^+^ subset was significantly depleted in the higher-stage N1 LNs. Lastly, GC B cells, plasmablasts (*CD38, MKI67*), and plasma cells were all observed to be more abundant in higher-stage N1 LNs compared to all other LNs, suggesting an increased rate of GC B cell differentiation into plasma cells in tumor-adjacent LNs of higher-stage patients (FDR < 0.05; **Fig. 2G**, **Supplementary Fig. 8**). Together, these results suggest that dysfunctional T-cell phenotypes, GC B cell differentiation into plasma cells, and depletion of NK cells are features of the non-metastatic LNs in patients with higher-stage NSCLC.

**Fig. 2.**
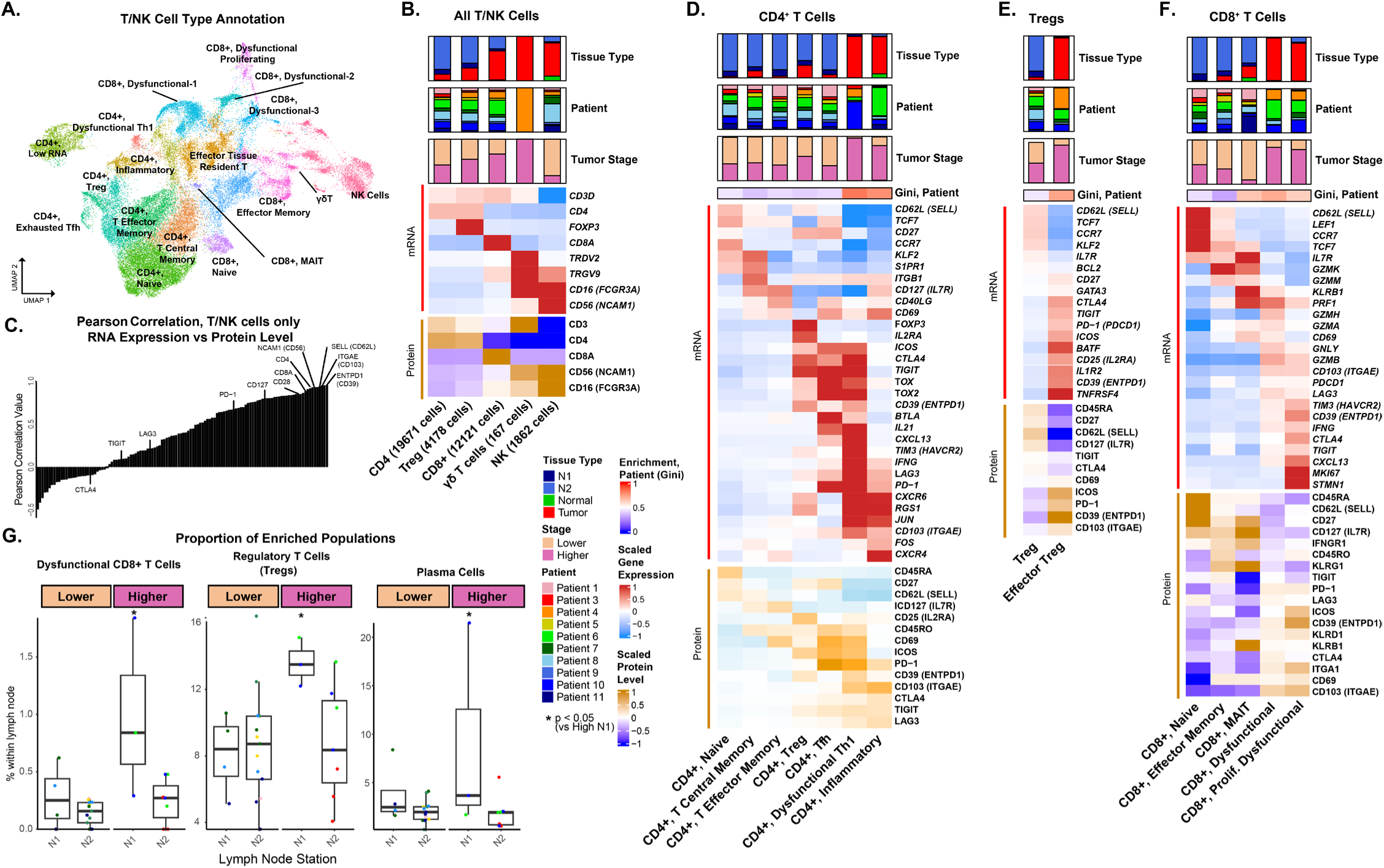
Deep phenotyping of lymphocyte populations with multimodal profiling revealed enrichment of regulatory T cells, and dysfunctional CD8^+^ T cells in N1 LNs of higher-stage patients. **A)** RNA expression-based UMAP projection of 47,068 T cells and NK cells, color-coded by cell type annotations. **B)** Heatmap of broad T cell/NK cell type annotations, categorized into distinct subgroups: non-regulatory CD4^+^ T cells, regulatory CD4^+^ T cells (Tregs), CD8^+^ T cells, γδ T cells, and NK cells. Relative proportions of each cell population observed across sampled tissue types, the originating patients and the tumor stages are represented in bar chart form. Gene (top) and protein (bottom) cell type specific markers are displayed in a heatmap. Each marker was Z-score normalized across all T/NK cells and averaged per subpopulation. CD4^+^ T cell subtypes were largely enriched in LNs whilst CD8^+^ T cells were observed more frequently amongst the tumor samples. The concordance of RNA expression and surface protein levels was assessed for 122 markers in the CITE-seq panel by examining the Pearson correlation between the average RNA and protein levels across cell populations. High correlations were observed for most broad cell type markers while some markers of T-cell activation displayed lower concordance. Heatmaps of selected RNA and protein markers are shown for **D)** CD4^+^, **E)** Treg, and **F)** CD8^+^ T cell subpopulations. All markers were differentially expressed for their respective subpopulation compared to all other T cells (FDR q-value < 0.05, Log2FoldChange > 0.25). Expression is scaled across T cell types and averaged per subpopulation. The Gini coefficient was used to assess the dispersion of cell subpopulation proportions across patients. A higher Gini score indicates that the proportions for cell subpopulation were higher in a subset of patients while a lower score indicates that the proportions were more evenly spread across patients. **G)** A logistic regression model was used to identify cell populations enriched or depleted in the N1 LNs of higher-stage patients while controlling for patient. Dysfunctional CD8^+^ T cells, regulatory T cells, and plasma cells were enriched in N1 LNs of higher-stage patients. *Abbreviations for cell populations: TCM – Central Memory T cells, Tfh – T-follicular helper cells, Th1 – T-helper type 1 cells, MAIT – Mucosal associated invariant T cells*.

### Deep multimodal phenotyping of myeloid cells identifies enrichment of cDC2-like CD1c^+^ mregDCs in the N1 LNs of higher-stage patients

To investigate subpopulations of myeloid origin, 17,392 myeloid cells (*CD14*^+^) and pDCs (*CD123*^+^) were subclustered to identify 19 distinct subpopulations (**Fig. 3A**; **Supplementary Fig. 15**). These subpopulations were categorized into 5 major cell types: alveolar macrophages (*MARCO*^+^, *FABP4*^+^, *CD163*^+^), macrophages (*CD163*^+^), monocytes (*FCN1*^+^, *S100A8*^+^), conventional dendritic cells (cDCs; CD1c^+^), and pDCs (*CD123*^+^) (**Fig. 3B**). While high correlation was observed between RNA and protein for some dendritic cell markers such as FCER1A, CD1c, and CD123 (R > 0.7), a lower correlation was observed for other important markers such as CD141 (R = 0.500), CD163 (R = 0.550), CD14 (R = 0.573), and CD11c (R = 0.351; **Supplementary Fig. 16**). The integration of surface protein levels was important for resolving macrophage and dendritic cell heterogeneity, as transcriptomic expression alone was insufficient to distinguish specific subsets such as cDC1s defined by protein levels of CD141. Macrophage subpopulations included those with various M2 and M1 phenotypes as well as TAM expressing *SPP1*, *VEGFA*, and *CXCL8* with reported roles in stromal-remodeling and immunosuppression^47^ (**Supplementary Fig. 17**; **Supplementary Tables 15, 16**). In addition to the macrophages, *CD16*^+^/*CD14*^low^ and *CD16*^-^/*CD14*^high^ monocytes were also identified. Most macrophage and monocyte populations were enriched in the tumor and normal lung tissue samples, except for one LN-enriched macrophage population (*APOE*^+^, *LYZ*^+^, *CCL18*^+^, *CCL3*^+^, *CD68*^+^, *CD163*^+^; FDR < 0.05).

Dendritic cells (DCs) serve multiple functions within the immune response, including antigen presentation for training the adaptive immunity. The six dendritic cell subpopulations included mature regulatory dendritic cells (mregDCs; *LAMP3*^+^, *PD-L1*^+^, *CCR7*^+^, *IDO1*^+^, *CD86*^+^), Langerhans cells (LCs; *CD207*^+^, *CD1A*^+^), conventional dendritic cell subset 2 (cDC2; CD1c^+^, *CD11c*^+^, *FLT3*^+^), conventional dendritic cell subset 1 (cDC1; *CD141*^+^, *CLEC9A*^+^, *FLT3*^+^), pDC (CD123^+^, CD45RA^+^), and proliferating DCs (*MKI67*^+^, *TOP2A*^+^) (**Fig. 3C**). Proliferating DCs and LCs both expressed TIM3 and were predominantly found in a tumor sample from a Stage IIIA patient who was also a current smoker. mregDCs are key contributors to immunosuppressive tumor microenvironments and have been associated with poor patient outcomes^48–50^. However, their roles within LNs remain underexplored— particularly in the context of their interactions with recently described dysfunctional T cell populations in the tumor tissue of NSCLC or in the LNs near Head & Neck cancers^51,52^. As mregDCs mature from cDC1 or cDC2, they lose expression of their original cell-type markers such as *CD1C*, *FCER1A*, *CD141*, and *CLEC9A* at the transcriptional level^53^. While we observed a general lack of RNA expression for *CD141* and *CD1C* in the mregDC subpopulation, higher levels of CD1c CITE-seq expression were detected in the majority of mregDCs (**Fig. 3D**). In contrast, the cDC1 subpopulation maintained relatively high surface protein levels of CD141 and low surface protein levels of CD1c compared to the cDC2 and mregDC subpopulations (**Fig. 3E**). This data suggests that mregDCs in our dataset may originate from cDC2s which specialize in antigen presentation to CD4^+^ T cells such as Tregs^28,49,53,54^. Additionally, an increased ratio of cDC2 to cDC1 has been associated with worse clinical outcomes in cancer patients^55,56^. In our cohort, the proportion of cDC2-like mregDCs was significantly enriched in the higher-stage N1 LNs compared to remaining LNs (FDR = 0.021; **Fig. 3F**). Moreover, the proportions of mregDCs were correlated with the proportions of dysfunctional CD8^+^ T cells and Tregs across the LNs suggesting they may be facilitating immunosuppression in the LNs by interacting with cells in the T cell-mediated immunity pathway (**Fig. 3G, H**).

**Fig. 3.**
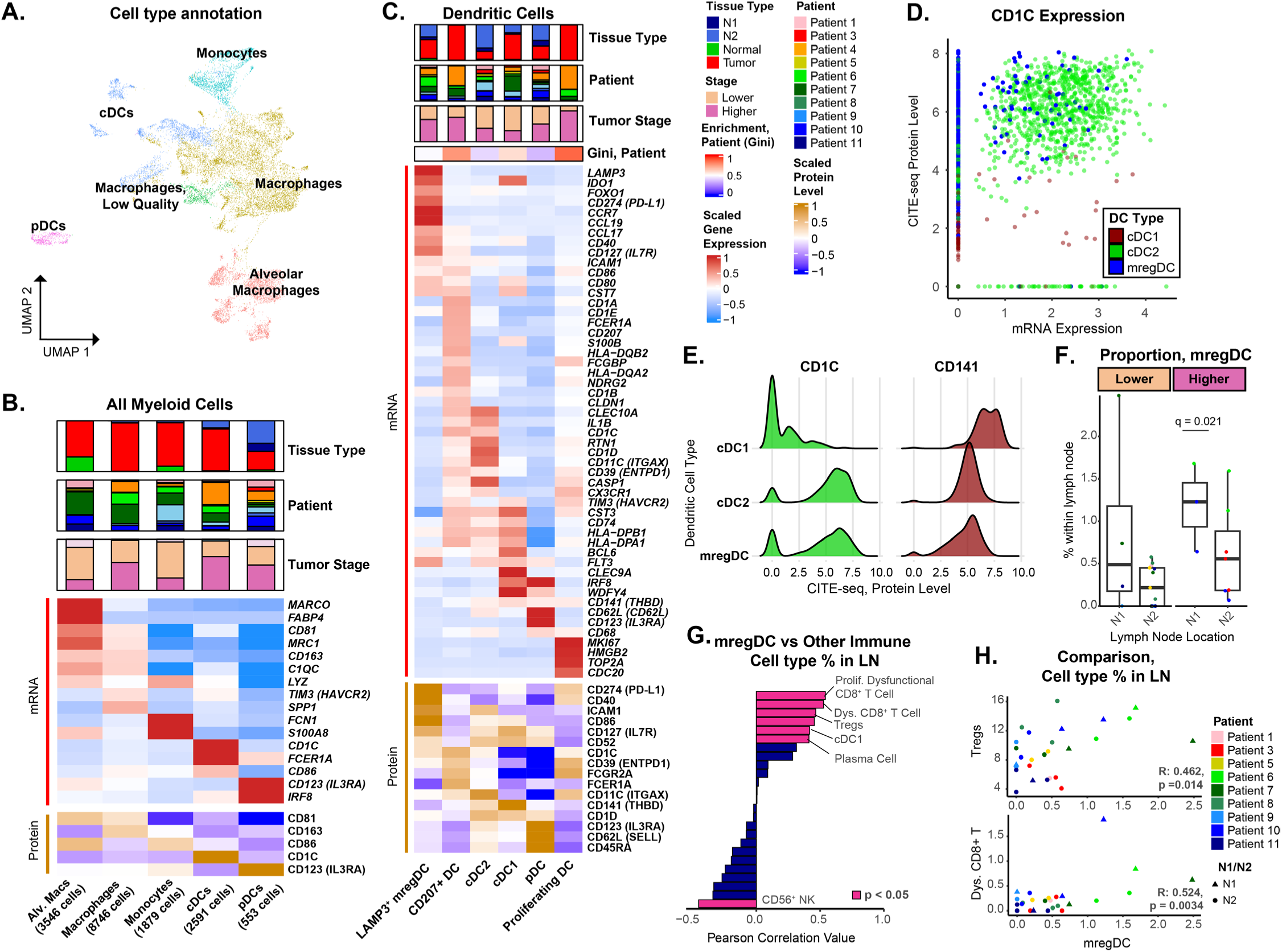
Multimodal profiling of myeloid cells identifies enrichment of cDC2-like mregDCs in the N1 LNs of higher-stage patients. **A**) RNA expression-based UMAP projection of 17,392 myeloid cells, color-coded by major cell type annotations. **B)** Average expressions of gene and protein markers are shown for the 5 major myeloid cell types. Expression for each marker was Z-score normalized across all myeloid cells and then averaged per major cell type. **C)** Differentially expressed gene and protein markers are shown for the six dendritic cell subpopulations (FDR < 0.05, Log2FoldChange > 0.25). Each marker was Z-score normalized across all DC cells and averaged per subpopulation. **D)** mRNA expression of *CD1C* (x-axis) is compared to the surface protein levels of CD1c (y-axis) for all DC subpopulations. CD1c is the primary marker for the cDC2 cells. The majority of mregDCs displayed low RNA expression of *CD1C* but high expression of CD1c on the protein level suggesting they are derived from cDCs in origin. **E)** The distribution of surface protein levels for CD1c (left) and CD141 (right) are shown for cDC1, cDC2, and mregDC subpopulations. The cDC2 and mregDC subpopulations expressed high levels of CD1c compared to the cDC1s subpopulation while CD141 was relatively higher in the cDC1s. **F)** The proportions of the mregDCs measured by scRNA-seq in the LNs are depicted. Each point represents a LN with colors indicating the respective patient from which the sample was collected. The proportion of mregDCs was significantly higher in the N1 LNs from higher-stage patients. **G)** The correlation between the proportion of mregDCs and each other immune cell population was calculated across LNs. Immunosuppressive immune cell populations such as dysfunctional CD8+ T cells and Tregs were significantly positively correlated with presence of mregDCs, while naïve B cells and NK cells were significantly negatively correlated. **H)** Scatterplots illustrating the positive correlation between the proportion of mregDCs and the proportion of Tregs (top) and dysfunctional CD8+ T cells (bottom) within the LNs. Each dot represents a single LN and is colored by patient. The shape of the dot denotes the nodal region. *Abbreviations for cell populations: Alv. Macs – Alveolar macrophages, cDCs - Classical dendritic cells, pDCs - plasmacytoid dendritic cells, mregDCs-mature regulatory dendritic cells*.

### Spatial analysis reveals altered immune spatial interactions in N1 LNs of higher-stage patients

To distinguish additional cell populations in the IMC data, unsupervised clustering was applied to each of the 8 major cell types and revealed a total of 50 distinct cell clusters (**Supplementary Fig. 18A**). Of particular interest were distinct IMC expression profiles of DCs (**Fig. 4A**), macrophages (**Fig. 4B**), and B cells (**Fig. 4C**). In general, the spatial distribution of different B-cell, T-cell, fibroblast, and myeloid clusters could be used to define specific regions within the LNs. B cell clusters corresponded to distinct regions of secondary B cell follicles. BCell-1 (Ki-67^+^) corresponded to the dark zone while clusters BCell-5 (CD138^+^, CD86^+^) and BCell-6 (CD138^+^, PD1^+^, CD45RO^+^) denoted the light zone (**Fig. 4D**). In stimulated B-cell follicles, clusters BCell-3 and BCell-4 largely defined the mantle zone. A layer of CD141^+^ fibroblastic reticular cells (FRCs) could be found throughout the cortical zones and often denoted the boundaries of B-cell follicles. LAMP3^+^ DCs, CD8^+^ T cells, and CD4^+^ T cells were predominantly present in the T cell zones in the paracortical regions.

We next examined different myeloid subpopulations and their association with nodal regions, stage, and LN architecture. Three distinct subpopulations of mregDCs were identified, including DC/mregDC-5 (LAMP3^+^, CD1c^+^, TIM3^+^, CD86^+^, PD-L1^+^), DC/mregDC-2 (LAMP3^+^, CD86^+^), and DC/mregDC-8 (LAMP3^+^, TIM3^low^). Additionally, cDC1 (DC/mregDC-4; CD1c^-^, CD141^+^), and cDC2 (DC/mregDC-7; CD1c^+^, CD141^-^) were also detected. Notably, the proportion of mregDC-5 was enriched in N1 LNs of higher-stage patients compared to all remaining LNs (FDR < 0.05; **Fig. 4E; Supplementary Figs. 19, 20**; **Supplementary Table 17**), concordant with the association identified in the CITE-seq data. Macrophage subpopulations included CD68^+^ (Macrophage-2), CD163^+^, TIM3^+^ (Macrophage-3,4), and CD14^+^, TIM3^+^ subsets (Macrophage-6) (**Supplementary Fig. 18**). Macrophage-2 was enriched in lower-stage N1 LNs (FDR q-value = 0.013), whereas Macrophage-4 (CD163^+^, TIM3^+^, CD86^+^) was more prevalent in higher-stage N1 and N2 LNs (FDR q-value = 0.004). When examining the spatial locations, we observed that the mregDC-5 cluster was predominantly outside of B cell follicles in both the H&E and IMC, while the Macrophage-4 cluster was localized within B cell follicles of the N1 LNs from higher-stage patients (**Fig. 4F**).

Due to the densely packed nature of the tissue and the characteristics of antibody staining, some clusters were defined by not only the protein expression of the native cell type but also by protein expression from spatially adjacent cell types. Consequently, these clusters inherently capture information about the spatial proximity to neighboring immune cell types in addition to the cluster’s native cell type. For instance, the BCell-6 cluster displayed a high expression of PD-1 and CD4 alongside GC markers CD38 and CD138 indicating presence of CD4^+^ Tfh cells neighboring the GC B cells (**Fig. 4C**). To rigorously quantify the number of potential interactions between all pairs of cell clusters, we calculated the average number of cells from one cluster that were within a 20μm radius of the other cell clusters within each ROI and determined if the average number of potential interactions for each pair was significantly different between groups of LNs. Inferred interactions between the Macrophage-4 cells and the germinal center B cells were found more often in the N1 LNs of higher-stage patients compared to all other LNs (**Fig. 4G, Supplementary Table 18, Supplementary Fig. 19B**). Additionally, a closer proximity was found between the mregDC-5 cells and the cells from the dysfunctional CD8-5 cluster (CD8^+^, TIM3^+^, PD-1^+^, CD45RO^+^, CD45RA^-^) suggesting that interactions with a subset of mregDCs can promote CD8 T cell dysfunction. TAMs have been shown to support tumor growth by suppressing immune cell cytotoxic function and facilitating immune evasion^57,58^. Surprisingly, we also found increased neighboring between TAM-3 and BCell-4, suggesting that increased interactions between the CD163+ TIM3^dim^ TAM population and mantle zone B cells occur in the regional LNs of patients with more advanced stage disease (**Fig. 4H**).

**Fig. 4.**
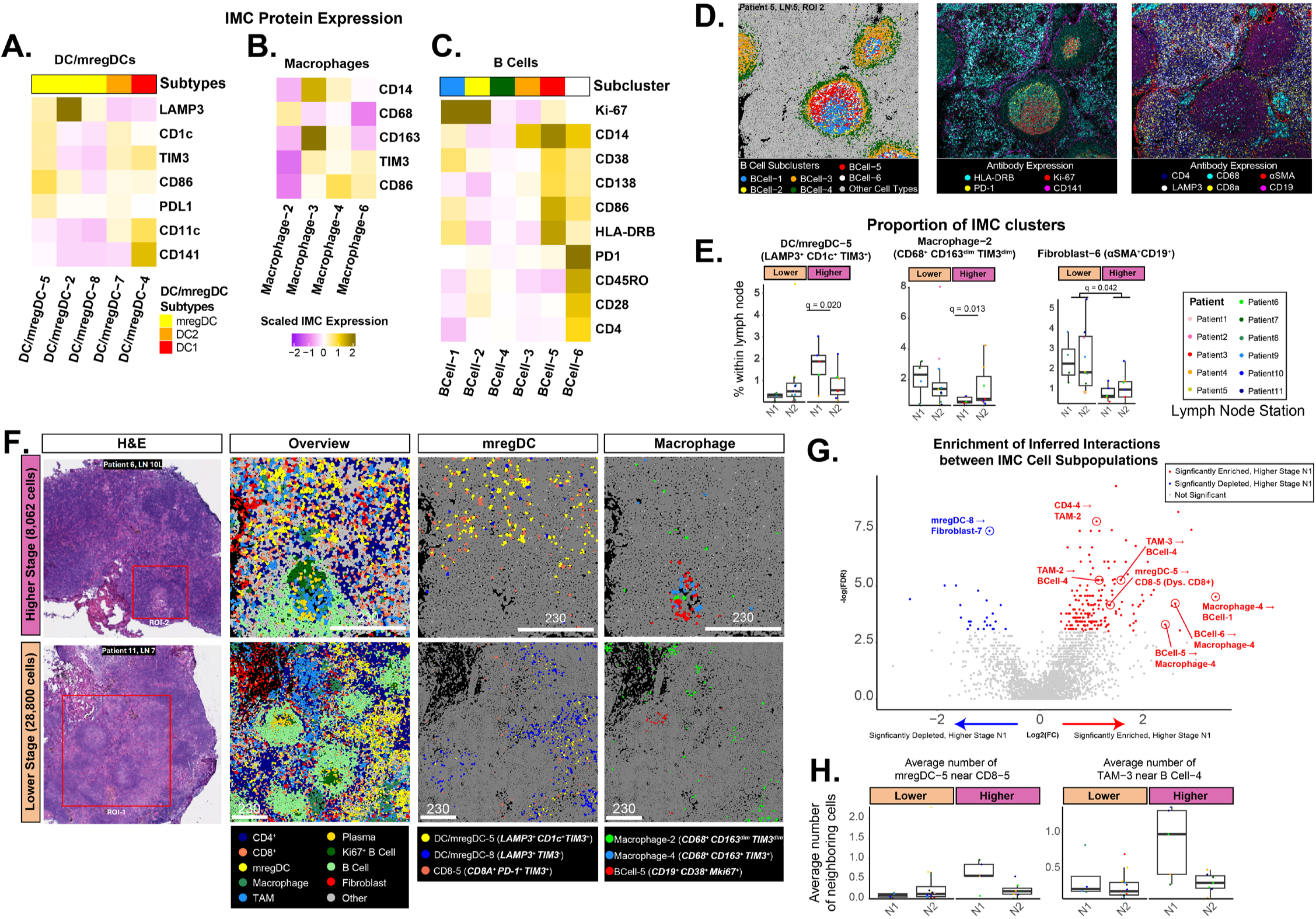
Imaging mass cytometry (IMC) reveals immune cell interactions enriched in the N1 LNs of higher-stage patients. Subclustering of IMC cell types revealed additional clusters of **A)** DC/mregDC, **B)** macrophages, and **C)** B cells. **D)** Different B cell clusters are spatially organized and reflect B cell follicle structure including the light, dark, and mantle zones. **E)** The proportions of the mregDC-5, Macrophage-2, and Fibroblast-6 clusters measured by IMC in the LNs are depicted. Each point represents a LN with colors indicating the respective patient from which the sample was collected. Macrophage-4 and mregDC-5 (LAMP3^+^, CD1c^+^, TIM3^+^, CD86^+^, PD-L1^+^) are more prevalent in LNs of higher-stage patients compared to LNs of lower-stage patients. Conversely, the Macrophage-2 and Fibroblast-6 (αSMA^+^, CD19^+^) clusters can be found more commonly in the other LNs. **F)** Spatial distributions of macrophage and mregDC clusters that are enriched or depleted in the N1 LNs of higher-stage patients. H&E is shown to denote ablated regions. Single cells are colored by overall cell type labels or specific macrophage, mregDC, and T-cell cluster labels. Macrophage-4 (CD163^+^, TIM3^+^, CD86^+^) and mregDC-5 (LAMP3^+^, CD1c^+^, TIM3^+^, CD86^+^, PD-L1^+^) are more prevalent in LNs of higher-stage patients compared to LNs of lower-stage patients. Conversely, the Macrophage-2 and mregDC-8 clusters can be found more commonly in LNs of lower-stage patients. *Lymph node samples shown: Higher-stage – Patient 6, LN 10L, ROI 2; Lower-stage – Patient 11, LN 7, ROI 1. Lower - Lower-stage patient, Higher - Higher-stage patient.* **G)** To quantify the number of cell-cell interactions between all pairs of cell clusters, the average number of cells from one cluster that were within a 20μm radius of the other cell cluster was calculated. A Wilcoxon-ranked sum test was then used to determine if the average number of interactions for each pair was significantly different between groups of LNs. **H)** The average number of interactions between the mregDC-5 and CD8-5 clusters (left) and between the TAM-3 and BCell-4 clusters (right) was significantly increasing in higher-stage N1 LNs.

### Spatial niche analysis reveals decorticated follicular architectures in the N1 LNs of higher-stage patients

To identify spatially coordinated immune microenvironments in LNs, we clustered cells into niches based on the cell subset marker profiles (neighbors) that were present within a 20 µm radius (**Methods**, **Fig. 5A**). The mean number of neighbors per cell was 17.2. This analysis identified 40 unique spatial niches across all LNs (**Supplementary Fig. 21, Supplementary Table 19**). Mapping these niches onto tissue sections revealed that active, secondary B cell follicles were primarily composed of Niches 23, 14, 10, 21, and 13 (**Fig. 5B**). Niche 23 was enriched for Ki-67⁺ BCell-1 and BCell-2 populations, as well as CD138⁺ plasmablasts, consistent with the GC (**Fig. 5C, Supplementary Fig. 22**). The mantle zone (MZ) structures, which encapsulate the GC, consisted of Niches 10, 13, 14, and 21. Dormant B cell follicles lacked Niche 23 and were composed exclusively of MZ-associated niches (**Supplementary Fig. 23A, B)**.

Several of the follicular niches were differentially abundant in higher-stage N1 LNs. Niches 28, 5, and 39 were enriched in higher-stage N1 LNs while Niches 2, 16, 21, and 24 were significantly depleted (FDR q-value < 0.05; **Fig. 5D, Supplementary Figs. 24, 25, Supplementary Table 20**). Niche 21 was rich in mantle zone B cells (BCell-4; CD19⁺ CD45RA⁺) and formed compact MZ structures in lower-stage LNs (**Fig. 5E**). Niches 6 and 24 generally surrounded the follicular regions and were depleted in the N1 LNs of higher-stage patients. Niche 6 predominantly contained BCell-4, CD4-1 (CD4⁺ CD45RO⁺), and FRC clusters reflecting normal B cell follicle structure (**Fig. 5F, Supplementary Figs. 24, 25**). Niche 24 was composed of BCell-4, CD4-1, and CD4-3 (CD4⁺, CD45RA^high^, CD39^high^) which is consistent with normal follicular helper T cell activity (**Supplementary Fig. 25**). In contrast, Niche 28 was enriched in higher-stage N1 LNs and contained high proportions of BCell-4, Tregs (cluster CD4-4), TAM-2 (CD68^+^, CD163^+^, TIM3^high^, CD141^high^), TAM-3 (CD68^+^, CD163^+^, TIM3^low^), and TAM-4 (CD163^+^, TIM3^+^) clusters (**Fig. 5E, F**). Niche 28 was largely distributed throughout the N1 LNs of higher-stage patients suggesting that interactions between Tregs and TAMs may contribute to dysregulated follicular phenotypes.

In addition to dormant/primary B cell follicles and active/secondary B cell follicles, we identified a third category of B-cell-like secondary follicles that had a GC-like Niche 23 core but lacked surrounding MZ niches (**Supplementary Fig. 23C**). We named these follicles “decorticated” (i.e. removal of the bark, rind, or husk) due to the lack of immune niches normally present in secondary follicles. To quantify the degree of mantle zone degradation surrounding follicles with GCs, we first computationally identified all GCs as those with densely interconnected cells from Niche 23 (**Supplementary Fig. 26**). To further investigate niche changes surrounding all B cell follicle structures, we computationally identified all follicles by identifying regions that have densely connected GC and MZ niches 23, 10, 13, 15, and 21 (**Fig. 5G**). We next calculated the percentage of cells from the MZ niches among all cells within a 20 µm radius from the GC cells (**Fig. 5G**) and found that it was significantly lower in the higher-stage N1 LNs (**Fig. 5H**). Additionally, the proportion of MZ niches out of all cells within a 20 µm radius from B cell follicles was significantly lower in higher-stage N1 LNs (**Fig. 5I**). We also determined which cell clusters were enriched in follicular regions of N1 LN from higher-stage patients. Macrophage-4 and TAMs 2, 3, and 4 were significantly enriched in the follicles of higher-stage N1 LNs, while MZ B cells (BCell-4) and CD4^+^ T helper cells (CD4-3) were significantly depleted (**Fig. 5J, Supplementary Fig. 27**). Finally, to determine whether the regions adjacent to follicles had altered immune composition, we determined whether niches within a 100 µm radius of the follicular regions were different across stages or nodal regions. Niche 28 remained significantly enriched in the regions surrounding the higher-stage N1 LNs (**Fig. 5K**). This supports a model where increased interaction between Tregs, CD163^+^ TIM3^+^ TAMs, and CD68+ CD163^+^ TIM3^+^ TAMs occur along with the decortication of secondary B-cell follicles in the regional LNs of patients with more advanced stage disease (**Fig. 5L**).

**Fig. 5.**
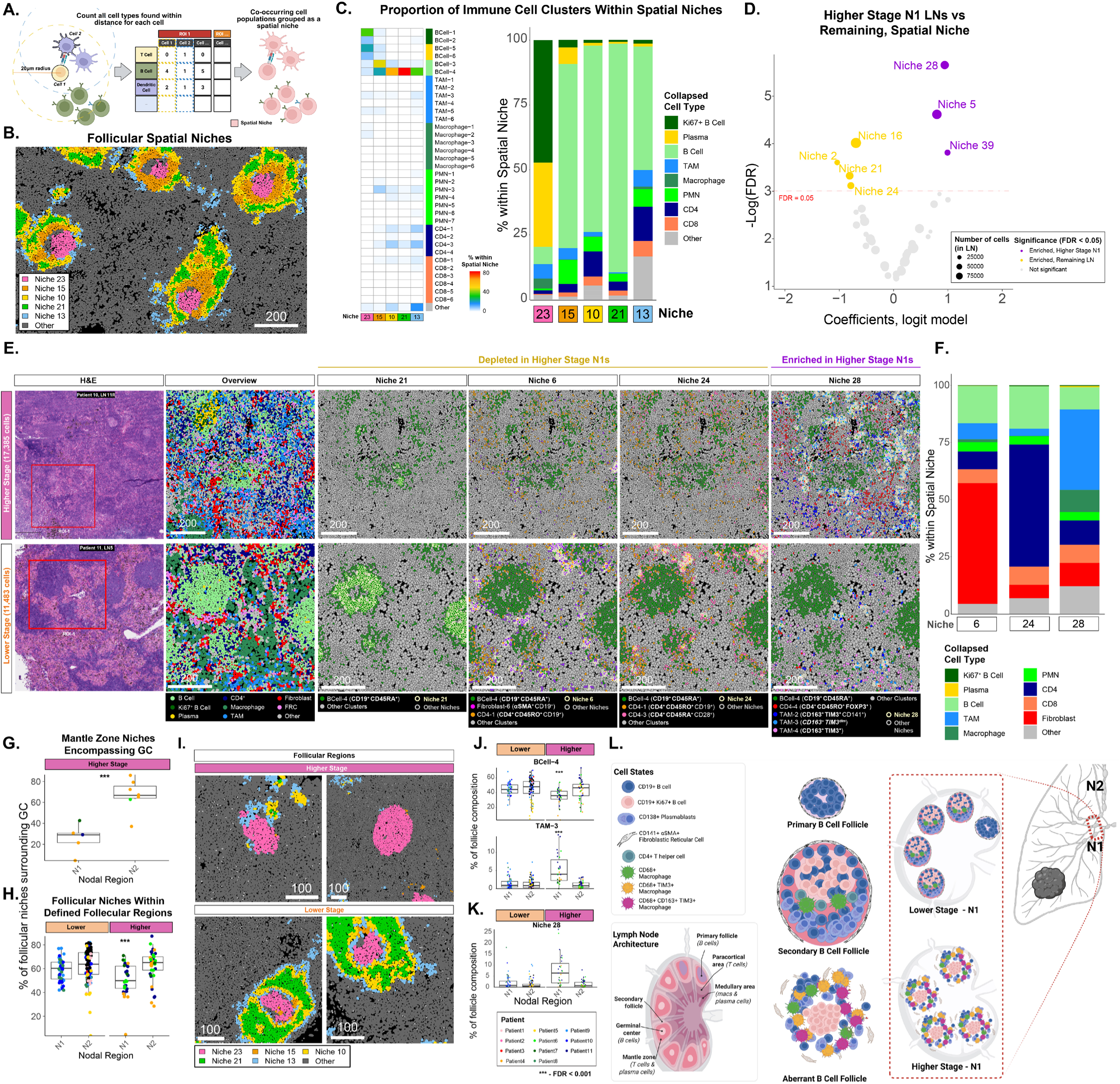
Altered cell composition and architecture of B cell follicles in N1 LNs of higher-stage patients. **A)** For each cell, neighboring cell labels within a 20µm radius were counted and then cells were clustered according to this neighborhood information to generate spatial niches. **B)** Masks for spatial niches predominantly located in B-cell follicular regions are colored including niches 10, 15, 21, and 23. All other cells classified as different niches are colored gray. **C)** Proportions of IMC clusters are shown for spatial niches involved in follicular regions (left). The proportions of the major immune cell types are shown for the same spatial niches (right). **D)** A logistic regression model was used to identify niches enriched in the N1 LNs of higher-stage patients. **E)** Example regions are shown for LNs from a lower-stage patient (Patient 11, LN 5, ROI 1) and a higher-stage patient (Patient 10, LN 10R, ROI 1). The red square in the H&E image denotes ROIs selected for ablation. An overview of the major cell types derived from IMC is shown in the second panel. The remaining panels show examples of niches enriched or depleted in N1 LNs from higher-stage patients. Cells within the niche have a thicker white border. Spatial Niche 21 contained high proportions of cluster BCell-4 and denoted follicular structures. Spatial Niche 6 contained clusters BCell-4 and Fibroblast-6 while Niche 24 contained BCell-4 and CD4-3 clusters. Both niches could be observed in the regions surrounding follicles. Spatial Niche 28 contained high proportions of cluster BCell-4, Tregs (cluster CD4-4) and several clusters of TAMs, and was distributed throughout the N1 LNs of higher-stage patients. **F)** Proportions of immune cell types composing the spatial niches 6, 24, and 28 are shown. **G)** Germinal centers were computationally defined as regions of interconnected cells from niche 23. Follicular regions were computationally defined as regions of interconnected cells from follicular niches 23, 10, 13, 15, and 21. Decorticated B cell follicles lacking mantle zones were more prevalent in the N1 LNs of higher-stage patients. Masks are colored using spatial niches depicting germinal centers (Spatial Niche 23), mantle zones (Spatial Niches 10, 13, 15, 21), and other cell populations. *Lymph node samples shown: Higher-stage – Patient 3, LN 10R, ROI 2; Lower-stage – Patient 9, LN 7, ROI 3.* **H)** The fraction of cells from mantle zone niches 10, 13, 15, and 21 was calculated in the regions surrounding GCs. In the decorticated follicles of higher-stage N1 LNs, mantle zone niches were less frequently observed in areas directly surrounding GCs. Each point is colored by patient. **I)** The fraction of the total follicle region occupied by mantle zone niches was calculated. The fraction of mantle zone niches was significantly lower within the follicle regions of higher-stage N1 LNs. **J)** The proportion of cells that occupy the follicular regions are shown for clusters BCell-4 (top) and the TAM-3 (bottom) which were decreasing in higher-stage N1 LNs. **K)** The proportion of cells that occupy the follicular regions is shown for niche 28 which was increasing in higher-stage N1 LNs. **L)** Graphical summary of different follicular architectures. Dormant and active B cell follicle structures have intact mantle zones. In contrast, decorticated B cell follicles exhibit an altered structure including a decrease in mantle zone niches surrounding a germinal center. These follicles also show an altered immune cell composition marked by an increase of TIM3^+^ TAMs.

### The immunosuppressive microenvironment in the N1 LNs of higher-stage patients is marked by co-localization of Tregs, mregDCs, and dysfunctional CD8^+^ T cells

In anti-tumor adaptive immunity, DCs internalize tumor antigens, migrate to regional LNs, and promote activation and proliferation of cytotoxic T cells^59–61^. In the scRNA-seq/CITE-seq data, a significantly increased proportion of dysfunctional CD8^+^ T cells (TIM3^+^, PD-1^+^), Tregs (CD4^+^, FOXP3^+^), and TIM3^+^ mregDCs was observed in higher-stage N1 LNs (**Fig. 2F**). When examining the IMC data, spatial niche 39 was primarily composed of mregDCs (34.7% of all cells within the niche), Tregs (9.1%), and dysfunctional CD8^+^ T cells (2.9%) and was also significantly enriched in the N1 LNs of higher-stage patients (FDR q-value = 0.022; **Fig. 6A, B**). This niche was predominantly localized outside of B cell follicles (**Fig. 6C**). Together, our data suggests that mregDCs (CD1c^+^, TIM3^+^, and LAMP3^+^) and CD4^+^ Tregs in the LN microenvironment are associated with dysfunctional CD8^+^ T cells expressing higher TIM3 and PD-1 protein levels. This interaction occurs more often in the non-metastatic N1 LNs of higher-stage patients and thus may contribute to the systemic immunosuppressive environment needed for cancer aggressiveness.

**Fig. 6.**
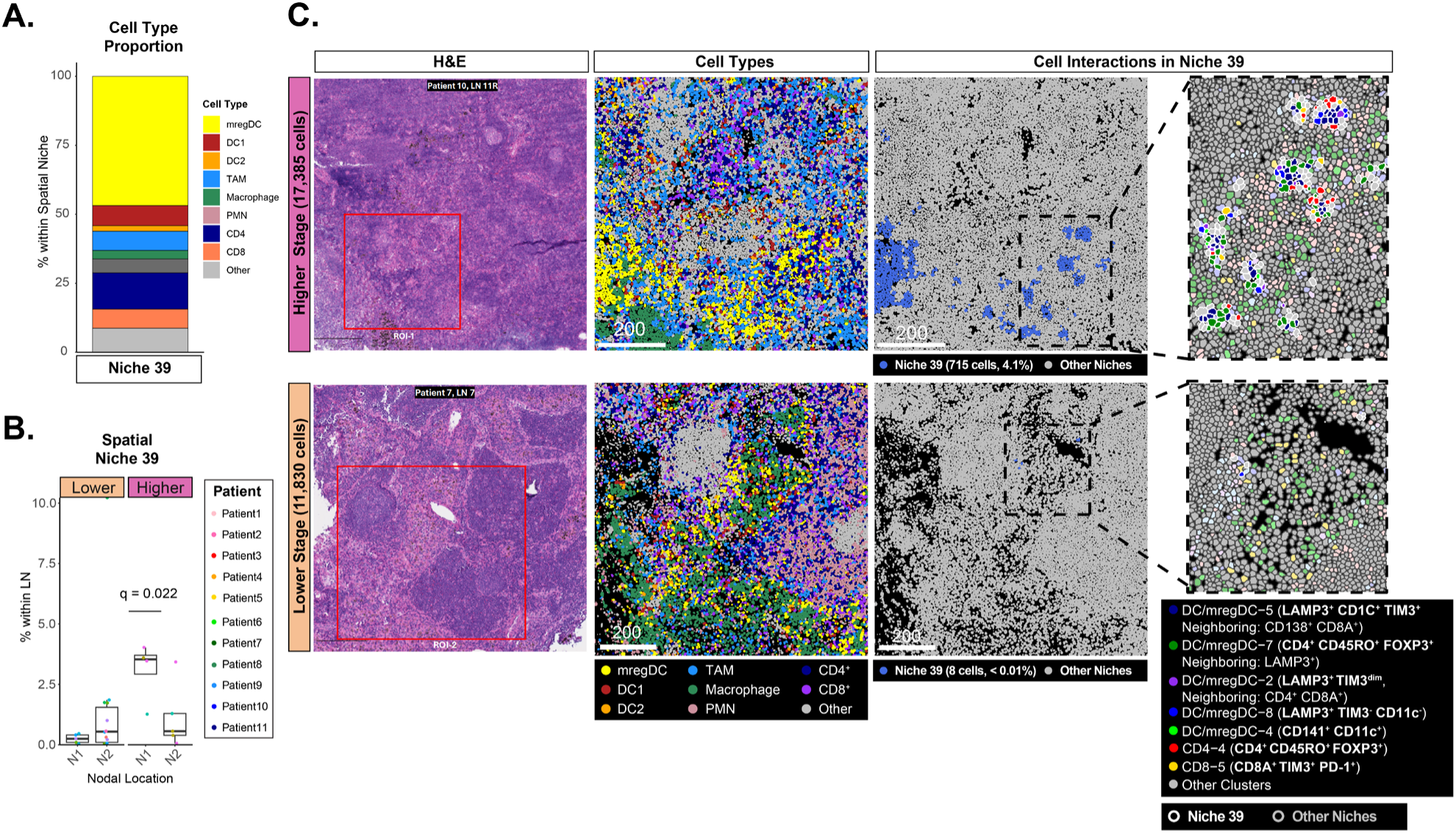
A niche of Tregs, mregDCs, and dysfunctional CD8^+^ T cells is enriched in the N1 LNs of higher-stage patients. **A)** Proportions of major immune cell types within Niche 39. mregDC-5 was the most predominant cluster in Niche 39. **B)** Niche 39 was significantly enriched in the higher-stage N1 LNs (FDR < 0.05). Each dot represents the proportion of Niche 39 within a LN and is colored by the patient from which the sample was obtained. **C)** Example regions are shown for LNs from a lower-stage patient (Patient 7, LN 7, ROI 1) and a higher-stage patient (Patient 10, LN 10R, ROI 1). The red square in the H&E image denotes ROIs selected for ablation. An overview of the major cell types derived from IMC is shown in the second panel. Cells within the niche have a thicker white border. Niche 39 is an immune neighborhood comprising clusters mregDC-5, mregDC-7, CD4-4 (Tregs) and CD8-5 (dysfunctional CD8^+^ T cells). In lower-stage LNs, Tregs and dysfunctional CD8^+^ T-cells are present but do not interact with mregDCs.

## DISCUSSION

Histologic characterization and genomic profiling of tumor tissue have become central to the diagnosis and treatment of NSCLC^62,63^. In contrast, clinical evaluation of the LNs remains limited to only the detection of the presence or absence of metastatic cells^64–67^. LNs are critical hubs for adaptive immune response, and their role in progression of NSCLC is a significant gap in our understanding of disease. Our study uses scRNA-seq, CITE-seq, and IMC to map the composition and spatial organization of immune niches in the non-metastatic LNs of patients with lower and higher-stage NSCLC. A comparison of hilar/peribronchial (N1) and mediastinal (N2) LNs from lower-stage (IA) versus higher-stage (IB–IIIA) patients identified two dominant immunopathologic axes of disruption: (1) dysregulation of humoral immunity marked by altered B cell follicular architecture and an increased immune spatial niche containing TIM3^+^, CD163^+^ macrophages within the surrounding immune microenvironment, and (2) altered cell-mediated immunity via an enriched suppressive immune niche containing dysfunctional CD8^+^ T cells, Tregs, and mregDCs.

Sentinel lymph nodes (SLNs) in breast cancer and melanoma represent the first anatomically predictable sites of metastasis. Pre-metastatic SLNs do not yet contain cancer cells but have been immunologically altered by the primary tumor to create an environment hospitable for future metastasis^20,68–76^. NSCLC may lack functionally equivalent SLNs due to its highly variable, multi-directional lymphatic drainage. Instead of being channeled through a defined serial path, lymphatic fluid from the lung often enters multiple intrapulmonary (N1) and mediastinal (N2) stations in parallel, consistent with the frequent occurrence of ‘skip’ metastases^77–81^. Our single-cell and spatial analyses reveal that non-metastatic N1 LNs of NSCLC patients undergo immune remodeling in higher-stage patients, particularly those with larger tumor burdens or concurrent metastasis in other nodes. While these immunosuppressive alterations may suggest a pre-metastatic niche similar to SLNs described in other cancer, they may instead reflect systemic changes associated with more advanced stage disease that are independent of the specific path of metastatic dissemination. Overall, these results underscore the need to evaluate regional LN immune states as key determinants of immune escape, progression, and therapeutic vulnerability.

Under homeostatic conditions, human B cell follicles in LNs are largely quiescent with intact architecture—characterized by round follicles supported by follicular dendritic cells and fibroblastic reticular cells (FRCs)^9,82,83^. Upon encountering foreign antigens, these follicles enlarge, and form germinal centers (GCs) comprising actively proliferating Ki-67^+^ B cells^84^. The germinal center is polarized into a dark zone for somatic hypermutation and a light zone for affinity selection, with unselected B cells cleared by macrophages^84–87^. Importantly, during activation, the GC is surrounded by a mantle zone, encapsulated by FRC, with follicles having defined follicular boundaries ^82,88,89^. However, deviations from traditional B cell follicle architecture have been pathologically described in cancer. In melanoma patients, B cell follicles in metastatic sentinel LNs have a less circumscribed architecture^90^. In the tumor-draining lymph nodes (TDLNs) of breast cancer patients, follicles were rounder in metastatic LNs and non-sentinel LNs compared to sentinel LNs, with the latter having non-circular germinal centers and macrophage infiltration in the cortex of the LNs^72^. Altered B cell follicle phenotypes have also been described in patients with common variable immunodeficiency (CVID) with irregular shaped and ill-defined GCs^91^, and with infections such as Salmonella that disrupt the LN’s B cell follicular architecture via LPS-elicited IFN-γ secretion from monocytes^92–94^. However, all previously reported architectures still maintained a predominantly intact mantle zone surrounding the germinal centers. Our IMC analysis reveals the presence of decorticated follicles enriched in the N1 LNs of higher-stage NSCLC patients. The term “decorticate” refers to the removal of the outer layer of bark or husk. Decorticated B cell follicles were characterized by loss of coordinated mantle zone niches, fragmentation of follicular boundaries, and increased plasma cell differentiation. Although further functional studies are required, the presence of decorticated B cell follicles may represent the loss of the development of effective or functional plasma cells in the humoral immunity response.

The decorticated follicles were associated with the presence of CD68^+^ CD163^+^ TIM3^+^ HLA-DR^high^ macrophages and CD163^+^ TIM3^+^ TAMs, rather than CD4^+^ T helper cells in lower-stage patients, suggesting a shift away from conventional T–B collaboration toward an immunosuppressive microenvironment. In contrast, low-stage N1 LNs and all N2 LNs retain well-demarcated primary and secondary follicles, with minimal CD163^+^ TIM3^+^ macrophage and TAM presence. The expansion of plasma B cells, whose prognostic role remains controversial, may reflect dysregulated humoral responses in higher-stage regional LNs, and may partially explain other’s findings of increased BCR clonotype diversity and fewer tumor-specific clonotypes in LUADs with increasing pathologic stage^95–99^. Furthermore, these results suggest that infiltration of CD163^+^ TIM3^+^ macrophages and TAMs may interfere with follicular structure and T-helper cells, compromising the LN’s ability to mount coordinated anti-tumor antibody responses. In solid tumor models, TAMs have been shown to impair CD8^+^ T cell proliferation and expand Tregs^17,74,76,100–106^. Previous studies have shown that patients with secondary B cell follicles in TDLNs are both associated with higher 5-year survival rate and better ICI response^107,108^. Our result raises the possibility that B cell follicle integrity and CD163^+^ TIM3^+^ TAM immune neighborhoods may serve as novel LN prognostic biomarkers for NSCLC patients. Previous studies in mice found that administering anti-TIM-3 antibodies during the early-stage LUAD significantly reduced disease progression, while administration at the later stages of disease did not have an effect and may explain the negative Phase III results for GSK’s cobolimab, an anti-TIM-3 mAb, for treatment of stage IV NSCLC^21,109–112^. Our data suggests that anti-TIM-3 treatments may be effective against metastatic spread in the regional LNs. Future mechanistic studies can investigate how TIM3^+^ CD163^+^ macrophages influence B cell follicle disruption, whether depletion of TIM3^+^ CD163^+^ macrophages will restore B cell follicle architecture, and how this shift in immune spatial architecture affects plasma cell clonality.

Parallel to this humoral alteration, we identified a distinct immunosuppressive niche involving Tregs, mregDCs, and dysfunctional CD8^+^ T cells. Specifically, we found that mregDCs—phenotypically like CD1c^+^ CD141^-^ DC2 subsets—frequently co-localized with FOXP3^+^ Tregs and dysfunctional CD8^+^ T cells in higher-stage N1 LNs. These CD8^+^ T cells exhibited elevated inhibitory markers consistent with reduced cytotoxic function. This spatial co-localization of regulatory populations suggests impaired priming and expansion of functional effector T cells within the LN. Prior studies in breast and colorectal cancers have similarly described enrichment of immunosuppressive elements, including Tregs, CD163^+^ macrophages, and dysfunctional DCs, in pre-metastatic LNs^20,21^. Other studies examining mouse models of lung cancer have found that regional LNs can harbor immunosuppressive cells, such as Tregs, to reduce CD8^+^ T cell cytotoxicity^75,113^. Our study extends these findings by spatially mapping the immunosuppressive niche and linking it to disease stage. TIM3+ mregDCs, Tregs, and exhausted CD8^+^ T cells coalesce into a structured suppressive microenvironment that may actively suppress cell-mediated immunity and enable tumor progression. These results suggest that novel immunotherapies which disrupt mregDC–Treg–CD8⁺ T cell interactions may restore T cell cytotoxic function and inhibit metastatic dissemination.

In our spatial analysis, although the LNs had dense cellularity, we were able to confidently assign each segmented cell to a major cell type based on the predominant marker intensity. However, some subclusters within a cell type exhibited expression of markers from other cell types. This likely did not represent a transition between cell types but rather “spillover” signal from the neighboring cells. This also explains why we obtained more clusters in the IMC data compared to scRNA-seq/CITE-seq despite having fewer markers. Despite its technical nature, this signal was still useful as it often reflected legitimate cell-cell interactions and spatial niches. In fact, several of the IMC clusters defined by other neighboring cell types were also found to be co-occurring in spatial niches. This challenge may be overcome in the future with the increasing specificity of signal intensities and better cell segmentation algorithms.

Our findings suggest several clinical implications for early-stage resected NSCLC. First, they support moving risk assessment beyond tumor size, nodal status, and tumor-cell PD-L1 expression alone to incorporate the immune architecture of both the primary tumor and regional lymph nodes. In current practice, pre-/peri-operative immunotherapy is indicated for resectable NSCLC with tumors ≥4 cm or node-positive disease in combination with platinum-containing chemotherapy, and adjuvant immunotherapy is indicated after resection for stage II–IIIA NSCLC with PD-L1 expression on ≥1% of tumor cells^39,114–117^. However, clinical outcome and response to immunotherapy can be heterogeneous, highlighting a need for prognostic as well as predictive markers of response to immunotherapy in this population^39^. Our findings suggest that an immunologically high-risk phenotype may already be detectable in morphologically uninvolved regional lymph nodes, defined by spatial niches enriched for dysfunctional or exhausted CD8+ T cells, Tregs, and mregDCs, in addition to decorticated B cell follicles. These features may identify a biologically higher-risk subgroup before overt nodal metastasis becomes evident, reflecting immune dysfunction despite complete resection. In the future, such patients could be prioritized for intensified postoperative surveillance, enrollment in adjuvant immunotherapy trials, or biomarker-directed escalation strategies. More broadly, our study suggests that lymph node immune microenvironment (LIME) may help refine recurrence-risk stratification beyond TNM stage and PD-L1 expression, although prospective validation will be required before these features can be used as stand-alone clinical decision tools. For clinical translation, a reduced FFPE-based mIF panel could provide a practical and cost-effective approach for detecting these spatial immune features in routine practice.

In conclusion, our multimodal and spatial profiling uncovers a dual axis of immune alterations. The immunosuppressive niches and disorganized B-cell follicular architecture in metastasis-free LNs from higher-stage patients may be useful as a prognostic biomarker to improve lung cancer staging. Given the relatively small patient numbers in our cohort, future studies will be needed to correlate these LN features with long-term clinical outcomes such as recurrence. Our results underscore the importance of spatial context in immune modulation. Immune-related biomarkers should consider not only cell-type abundance but also their spatial arrangements within the LN microenvironment. Future clinical and pathological assessment of LNs can include cost-effective immunohistochemical stains to assess LN functional immune state and abundance of decorticated follicles for enhanced cancer staging. Filling in these gaps could yield early biomarkers for relapse risk or efficacy of immunotherapy, unveil new neoadjuvant or intra-nodal immunotherapy targets, and tailor surgical approach to lymphadenectomy.

## METHODS

### Patient cohort and sample collection

Samples of LNs, primary tumors, and normal lung tissue were collected from surgical specimens of patients undergoing curative-intent resection at Boston Medical Center (Boston, MA). Mediastinal LN dissection was performed as per the Commission on Cancer guideline^118^, removing, when possible, 3 N2 station LNs and at least 1 N1 station LN. Pathologic stage was determined per 8^th^ edition of TNM Staging. Patients were classified as having a “higher” tumor stage based on a pathological stage of IB or higher (Patients 2, 3, 5, 9). Based on histopathological examination, one patient was diagnosed with benign hamartoma (Patient 1). The remaining stage IA patients were classified as “lower” tumor stage (Patients 4, 6, 7, 8, 10). The Institutional Review Boards (IRB) at Boston University Medical Center approved the study, and all subjects provided written informed consent.

### scRNA-seq and CITE-seq data generation

Tissues were immediately transported, dissociated into single cells, and processed into cDNA on the same day. To enrich for immune cells, tissues were gently minced and ground through a 70 μm nylon mesh strainer (70 μm Falcon™ Cell Strainers, Fisher Scientific, Waltham, MA, USA; Cat: 08-771-2) to obtain a single-cell suspension in RPMI medium (RPMI 1640 Medium, Gibco™, Thermo Fisher Scientific, Waltham, MA, USA). The cell suspension was transferred into 50 mL conical tubes and concentrated by centrifugation (∼300 × g, 8 min, 4 °C). The suspension was treated with RBC lysis buffer (RBC Lysis Buffer (10X), BioLegend, San Diego, CA, USA; Cat: 420301), followed by the removal of dead cells using the Dead Cell Removal Kit (Miltenyi Biotec, Bergisch Gladbach, Germany; Cat: 130-090-101). Cells were washed with DPBS (DPBS, no calcium, no magnesium, Gibco™, Thermo Fisher Scientific, Waltham, MA, USA; Cat: 14190250), concentrated by centrifugation (∼300 × g, 8 min, 4 °C), and blocked for 30 min using Human TruStain FcX™ Fc Receptor Blocking Solution (BioLegend, San Diego, CA, USA; Cat: 422302). Cells were counted, targeting a total of 500,000 cells. Hashtag antibodies (TotalSeq™-C anti-human Hashtag antibodies (5 to 15), BioLegend, San Diego, CA, USA) were used at a 1:10 dilution, and CITE-seq antibodies (TotalSeq™-C Human Universal Cocktail V1.0, BioLegend, San Diego, CA, USA; Cat: 399905) were diluted according to the manufacturer’s instructions. Incubation was carried out for 30 min on ice, with gentle pipette mixing every 10 min to prevent cell precipitation. Cell suspensions were then washed, counted, pooled, concentrated, and subjected to 5′ single-cell RNA sequencing using the Chromium Next GEM Single Cell 5′ Kit v2 and Chromium Next GEM Chip K Single Cell Kit (10x Genomics, Pleasanton, CA, USA), targeting a recovery of 10,000 cells using manufacturer’s protocols. Sequencing libraries were prepared using the Library Construction Kit (10x Genomics, Pleasanton, CA, USA) and sequenced on an Illumina NextSeq 2000.

### Preprocessing of scRNA-seq and CITE-seq data

Transcriptomic reads were aligned to the GRCh38-1.2.0 reference genome and quantified using CellRanger (v6.1.2, 10X Genomics, Pleasanton, CA). CITE-seq ADT and HTO reads were examined for antibody-oligonucleotide conjugate specific sequence barcodes through CellRanger’s feature indexing function. Demultiplexing of HTO-multiplexed samples was achieved through the HTODemux function from the Seurat package (v4.3.0) in the R programming language (v4.2.1)^119^. Singlets were determined as cell barcodes that express a singular type of hashtag oligonucleotide and were retained for downstream analysis. As described previously, a population of cells with a low mRNA expression and high surface CITE-seq expression characterized as “spongelets” are discerned through CITE-seq^120^. As such, barcodes annotated as an “empty droplet” by the CellRanger algorithm with a 99th percentile total CITE-seq expression were re-integrated into the dataset. All other barcodes labeled as “empty droplets” were removed.

### Cell quality control and filtering

The SCTK-QC quality control pipeline from the SingleCellTK package (v2.8.1) was applied to the transcriptomic expression data to generate RNA-seq quality control metrics including RNA-seq library size, mitochondrial gene expression and doublet prediction scores^121^. Cells with high mitochondrial gene expression (>15% of entire gene expression counts), indicating cell stress during tissue dissociation, were subsequently filtered out. Decontamination of ambient RNA contamination present in the mRNA expression data was conducted through the decontX R package^38,122^. All parameters were set at default values, and the unfiltered droplet/raw matrix was passed into the algorithm with the background parameter. Cells displaying a high level of ambient RNA contamination (> 0.9) were removed. Lastly, cells expressing less than 50 antibodies out of the 128-antibody panel were also discarded. The resulting 88,203 cells (97.2% of 90,694 cells) were used for downstream clustering and analysis.

### Cell clustering and annotation with scRNA-seq and CITE-seq data

The decontaminated mRNA expression data was log transformed, and the expression of the top 2,000 features with the highest variability was scaled and centered. Linear dimensional reduction (PCA) was conducted, and a K-nearest neighborhood graph was derived from the 1st to 30th PC dimensions. The data was clustered at a resolution parameter of 1.0 through the Louvain algorithm in the Seurat FindClusters function. Through expression of canonical marker genes and surface protein markers, each cluster was then annotated broadly as one of the following: T/NK cell, B cell, plasma cell, pDC, other myeloid cells, neutrophil, mast cell, and epithelial cell. (Canonical marker gene: T cell/NK cell – CD3D, CD56, NKG7, B cell – CD19, MS4A1, plasma cell – CD138, MZB1, pDC – CD123, myeloid – CD14, CD163, neutrophilS100A8, mast cell – TPSAB1, epithelial cell – EPCAM; Canonical surface protein marker: T/NK cell– CD3D, CD56, B cell – CD19, CD20, plasma cell – CD38, pDC – CD123, myeloid – CD14, neutrophil-CD16, mast cell – CD117, CD82). Each broad cell population was subset and the aforementioned clustering procedure was repeated. Cell clusters displaying hybrid expression of the canonical marker genes from other cell types in the subclustering analysis were removed due to suspicion of doublets. In total, 85,869 out of 88,203 (97.4%) cells were used in the subclustering analysis. The clustering procedure was repeated once more at a resolution parameter of 0.8 to generate clusters taken for downstream analysis. Cell clusters were annotated based on differential expression of marker genes and proteins identified through the Seurat differential expression function FindAllMarkers (FDR q-value < 0.05, Log2FoldChange > 0.25). Within the function, differential expression of genes was conducted through applying the MAST generalized linear model controlling for the originating sample. The Wilcoxon rank sum test was applied for the differential expression of surface proteins. Cluster pairs of CD4⁺ T cells, B cells, and dendritic cells showing expression differences limited to X-linked, mitochondrial, ribosomal, or other housekeeping genes (e.g., MALAT1, MTRNR2L8, NEAT1), but otherwise exhibiting highly similar transcriptional profiles, were combined, resulting in reduction of 6 clusters to 3.

### Decontamination of ambient protein levels

ADT expression data was decontaminated through the DecontPro decontamination algorithm available within the decontX R package. The cell_type parameter was set to the mRNA expression-derived cell type clusters obtained through the aforementioned procedures. The prior variance for the ambient and background contamination level was set to 2E-5 and 2E-6, respectively.

### Associating cell subpopulations with patient-level phenotypes

The binomial logistic regression model from the sccomp package (v1.2.1) was applied to determine significant enrichment of cell populations in collected tissue type, tumor stage, and nodal regions. To focus on metastasis-free LNs, an LN with metastasis was excluded from this analysis (Patient 3, LN12R). An FDR q-value less than 0.05 was considered significant. Cells originating from stations 10L, 11L, 10R, 11R, and 12R were considered hilar origin (N1), while all other cells were considered mediastinal origin (N2). N1 and N2 nodes were treated indistinguishably when computing enrichment between LNs and tumors. Patient identification numbers were included as a random variable within the model. 6 patients, including Patient 1 (benign hamartoma), were grouped amongst lower-stage tumors. Another model was also run which compared N1 LNs from higher-stage patients with all other remaining LNs

### H&E staining, multiplex immunofluorescence, and image analysis

Hematoxylin and eosin (H&E) staining was performed using the Leica Autostainer XL, and slides were mounted with Leica Micromount (catalog number 3801730; Leica Microsystems, Wetzlar, Germany). Immunofluorescence (mIF) was carried out on a Ventana Discovery Ultra autostainer (Roche Diagnostics, Indianapolis, IN, USA). Pretreatment was performed with Benchmark Ultra CC1, a Tris-based antigen retrieval buffer, at 95 °C for 64 min. The secondary antibody used for all targets was Vector ImmPress™ Goat Anti-Rabbit IgG (catalog number MP-7451; Vector Laboratories, Newark, CA, USA), a prediluted solution incubated for 20 min at 37 °C following a protein-blocking step with Akoya Opal Diluent/Block (catalog number ARD1001EA; Akoya Biosciences, Marlborough, MA, USA). Slides were counterstained with Akoya Spectra DAPI (catalog number FP1490; Akoya Biosciences, Marlborough, MA, USA) and mounted using Prolong Gold Antifade Mountant (catalog number P36930; Invitrogen, Waltham, MA, USA). A summary of the assay, including information on primary antibodies, incubation times, fluorophores used, and the sequence of application, is provided in **Supplementary Table 3**. Whole slide-images were acquired with a PhenoImager HT^TM^ Automated Quantitative Pathology Imaging System and spectrally unmixed using InForm (v3.0).

### Imaging mass cytometry data generation

FFPE blocks of patient LNs and tumors were sectioned at 5 μm thickness and baked for 2 hours at 60 °C. FFPE slides were deparaffinized in xylene and rehydrated sequentially in 100%, 95%, 75%, and 40% ethanol for 5 min per step, followed by rehydration in ddH₂O for 5 min. Antigen retrieval was performed using antigen retrieval buffer consisting of pH 9 Tris-EDTA buffer (100x Tris-EDTA Buffer, pH 9.0, catalog number ab93684; Abcam, Cambridge, UK) with 1% glycerol (catalog number 911046-1L; Sigma-Aldrich, St. Louis, MO, USA). The antigen retrieval buffer was microwaved to boiling prior to submerging the tissue slides and continuously heated at 96 °C for 10 min. Slides were then cooled at room temperature for 30 min. Slides were washed twice with DPBS (DPBS, no calcium, no magnesium, Gibco™, Thermo Fisher Scientific, Waltham, MA, USA; catalog number 14190250). Tissues were blocked using 3% BSA and 1:200 Human TruStain FcX™ Fc Receptor Blocking Solution (catalog number 422302; BioLegend, San Diego, CA, USA) in DPBS for 30 min at room temperature. Tissues were stained with metal-conjugated antibodies diluted in DPBS containing 1% BSA and 0.1% Tween-20 at 4 °C overnight. All antibodies were previously conjugated to metal isotopes using Maxpar Antibody Labeling Kits (Standard BioTools, South San Francisco, CA, USA) following the manufacturer’s instructions. Details of the antibodies and conjugated metals used in the IMC panel are provided in **Supplementary Table 3**. Slides were then washed twice with 0.1% Tween-20 in DPBS and counterstained with Cell-ID Intercalator-Ir (Standard BioTools, South San Francisco, CA, USA) in PBS for 30 min at room temperature. Slides were washed with ddH₂O and air-dried. Ablation was performed at 1 μm resolution and 200 Hz using the Hyperion Imaging System (Standard BioTools, South San Francisco, CA, USA).

### IMC image processing and segmentation

Raw pixel intensity values were preprocessed prior to single-cell segmentation. Specifically, for each image file (ROI), the intensity values from all 40 protein channels were summed for each pixel. For pixels with summed intensities above the 99.99th percentile threshold, all channel intensities were set to zero. Additionally, pixels with intensities above the 99.99th percentile in two or more specified protein channels were zeroed out to address potential technical artifacts. For the 38 protein channels excluding nuclear channels (Ir191, Ir193) and the MaxPar IMC Cell Segmentation Kit (Pt196), the median intensity across all images was subtracted from each pixel.

Finally, to account for outliers, protein channel intensities were normalized to a 0–100 range using the 99th percentile of each marker. Single cell segmentation and intensity measurements were performed using the Steinbock toolkit (v0.14.1). Briefly, the Mesmer algorithm within the DeepCell library was employed to segment single cells from image files. The MaxPar IMC Cell Segmentation Kit, along with the CD45RA, CD45RO, and HLA-DRB channels, were used to define the cytoplasmic regions, while the two nuclear channels were used to delineate nuclei. Surface protein levels for each cell were calculated by averaging the pixel intensities within each segmented object for every channel. Protein level values were stored into a SpatialExperiment object through the IMCRtools (v1.4.2) read_steinbock function in the R programming language (v4.2.1).

### Association analysis of image-based single cell clusters

A two-step clustering approach was used to generate single-cell clusters from IMC data. Of the 35 antibodies included in the IMC panel targeting cell surface proteins, EpCAM and LAG3 were excluded due to high background signal. The Bayesian multinomial clustering model ‘celda_C’ from the Celda package (v1.14.2) was then applied to classify all cells into 8 broad cell types based on expression of the remaining 33 protein markers. The rate of perplexity change (RPC) was used to identify the optimal number of clusters. Cell clusters were manually annotated based on marker expression. (Protein Markers: Fibroblast – α-SMA; CD4^+^ – CD4; CD8^+^ – CD8A; B Cell – CD19^+^; PMN – CD14-, CD15^+^, CD68^-^; Macrophage – CD14^+^, CD68^+^; DC/mregDC – LAMP3^+^). Each major cell type was subsetted and the clustering procedure with the celda_C was repeated. All clusters were labeled by their primary cell type and secondary cluster label (e.g., Fibroblast-1). The same sccomp model described above was used to identify clusters associated with nodal location and tumor stage. Cells from all ROIs within a LN were treated as a single sample.

### Spatial assessment of cellular neighborhoods and cell niches

In each image, cellular neighborhoods were quantified based on the number of neighboring cells located within a 20-pixel radius. The neighborhood information was additionally used as input for K-means clustering to identify a total of 50 spatial niches. The total within-clusters sum of squares (WCSS) value was calculated to determine the optimal number of clusters. A Wilcoxon rank-sum test was performed to assess statistically significant proximity between two cell populations. Visual validation of these neighborhoods was conducted by examining each ROI using the MCDViewer software. Association of the spatial niche was conducted using the *sccomp* model described above.

## Code Availability

All code used to generate the analysis is available at https://github.com/campbio/Manuscripts/Xi_Lung_CITE_Seq.

## Data Availability

Single cell RNA-seq (scRNA-seq) and Total-seq/CITE-seq data is available in the Gene Expression Omnibus under accession GSE316782. Imaging Mass Cytometry data with corresponding images are available at Zenodo (https://zenodo.org/records/18265339).

## Author Contributions

SAM, KS, and JDC obtained funding for the project. KS identified patients and performed the surgical procedures, SM consented patients, EJB provided pathologic review of the biospecimens. ZHX generated the data with guidance from SAM and RP. YK and ZHX processed the raw data and performed analyses. YK and ZHX created the figures and wrote the manuscript. EEK supported data sharing, EB, PRH, JEB, EJB, SAM, KS and JDC edited the manuscript.

## Acknowledgments

This work was made possible by the support of Boston University’s Single Cell Sequencing Core, Comparative Pathology Laboratory, and Integrated Biomedical Imaging Suite.

