## Supplemental Figures for "Multimodal single-cell spatial profiling reveals altered T cell immunity and B-cell follicular architecture in non-metastatic lymph nodes of advanced NSCLC patients"

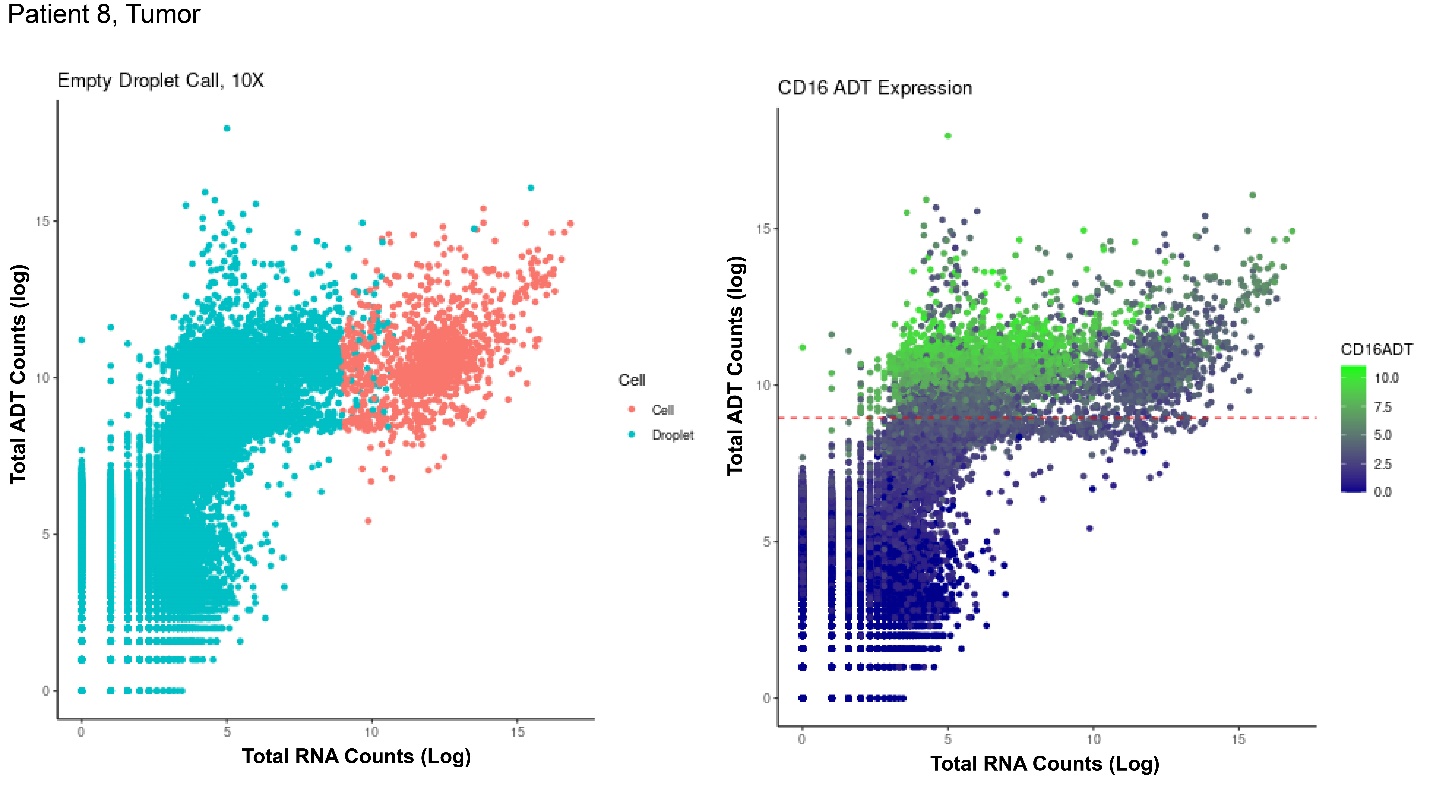

**Supplementary Figure 1. Recovery of neutrophil populations through CITE-seq expression.** Log-transformed total gene expression (x-axis) and ADT expression (y-axis) for all droplets from Patient 8’s tumor sample are shown. Droplets are color-coded by the empty droplet call generated by the 10X CellRanger algorithm (left) and the ADT expression level of the neutrophil marker CD16 (right). Empty droplets were classified as putative neutrophils and included in downstream analysis for each patient if they met all of the following criteria: (1) CD16 ADT expression above the mean, (2) total ADT expression more than two standard deviations above the mean, and (3) log-transformed gene expression greater than 5.

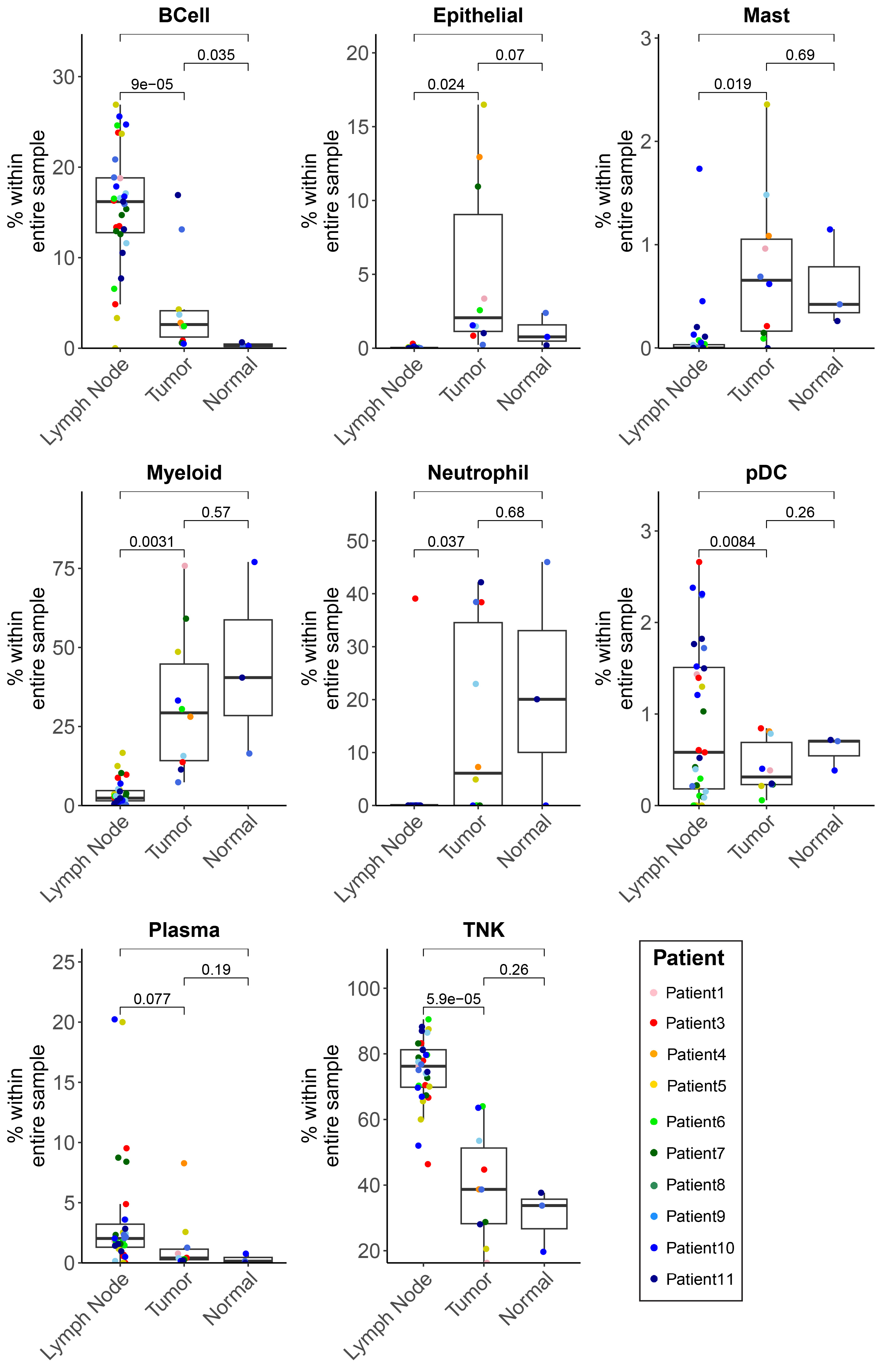

**Supplementary Figure 2. Comparison of cell type proportions across tissue samples.**  Boxplots display the proportion of each broad cell population derived from the scRNA-seq/CITE-seq data per tissue type. Each dot represents a sample, color-coded by the originating patient. The proportion of T and B lymphocytes was significantly higher in lymph node samples, while the proportions of myeloid cells and epithelial cells was significantly higher in tumor samples.

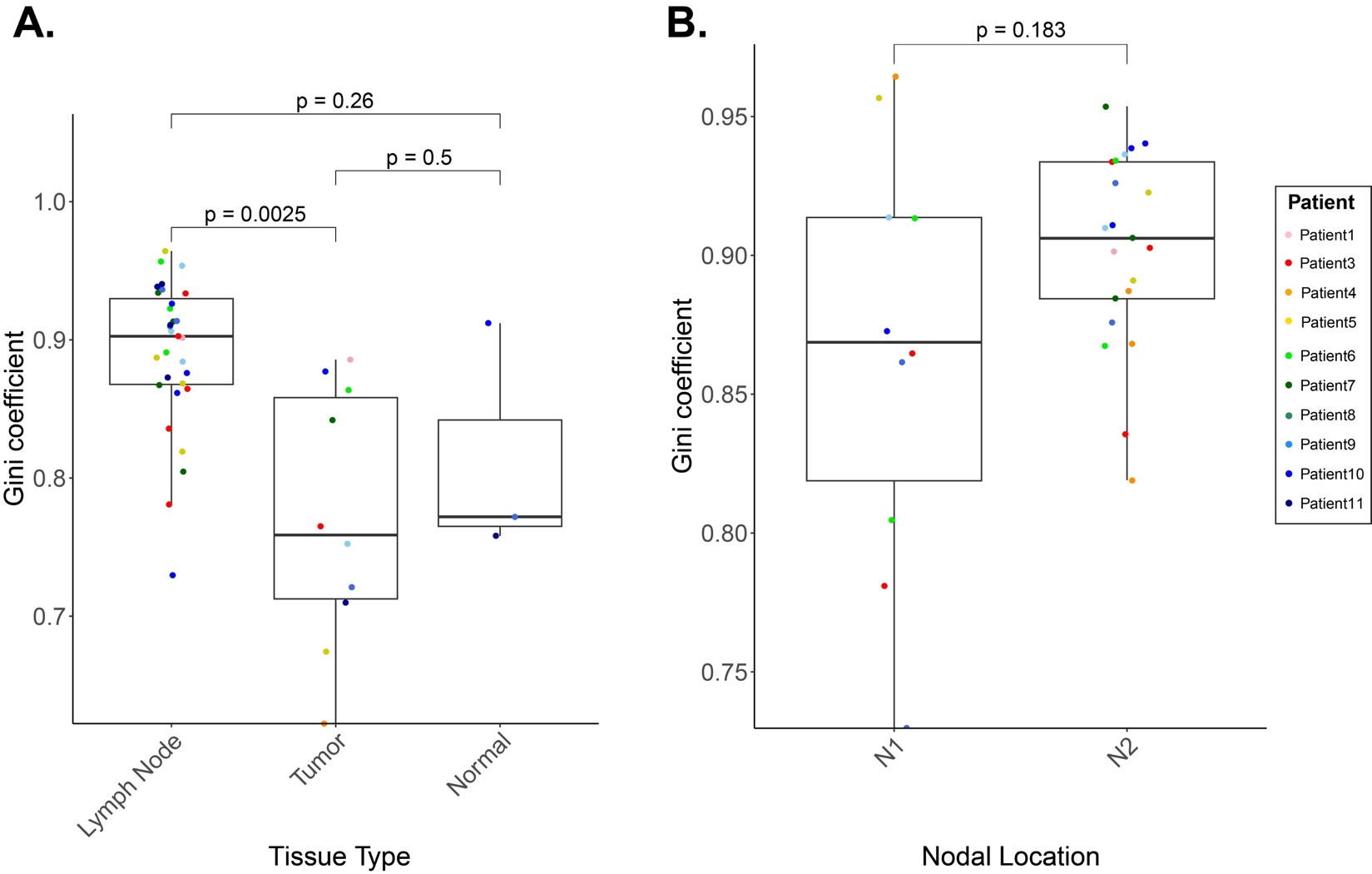

**Supplementary Figure 3. Assessing variability of cell type compositions from the scRNA-seq/CITE data across tissue types and nodal regions. A)** To measure the variability in cell type composition derived by scRNA-seq/CITE-seq, we calculated the Gini coefficient for each sample based on the proportions of the eight identified broad cell types (T/NK, B, plasma, myeloid, pDC, neutrophil, mast cell, and epithelial). A lower Gini coefficient indicates less variability of the cell types within the sample. Each dot represents a lymph node and is color-coded by the originating patient. On average, the lymph node samples exhibited significantly higher variability in cell type composition compared to tumor and normal lung tissue samples. **B)** The lymph node samples were additionally separated by nodal location (N1 or N2) to determine if variability of cell type composition differed by nodal location. No statistically significant differences were observed in the variability between broad cell types across the two nodal locations. P-values were computed using a t-test between each pair of tissue types or nodal regions.

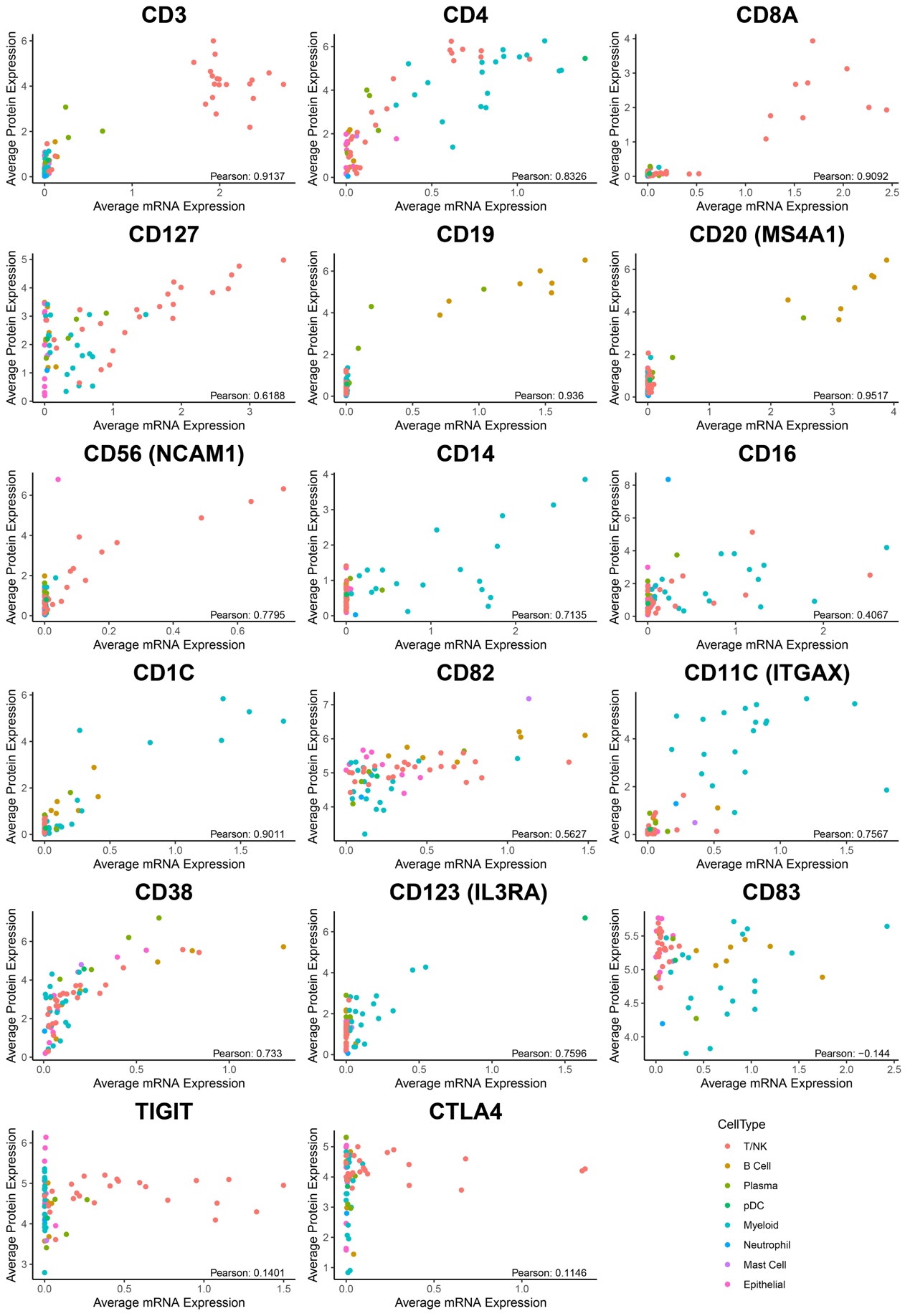

**Supplementary Figure 4. Correlation of RNA and corresponding protein expression for common cell type markers and activation states.**  Each of the eight major cell types (T/NK, B cell, plasma, pDC, myeloid, neutrophil, mast cell, epithelial) was reclustered, yielding a total of 66 subpopulations across all cell types in the scRNA-seq/CITE-seq data. Scatterplots display the average mRNA (x-axis) and corresponding protein (y-axis) expression levels of selected markers across the 66 subpopulations. Each dot represents the average expression for cell subpopulation and is color-coded by the broad cell type. The correlation was determined using a Pearson correlation coefficient. Canonical genomic markers for immune cells (ex. CD3, CD19, CD20, CD14) exhibited high concordance (R > 0.7) while markers associated with T lymphocyte exhaustion exhibited low concordance (R < 0.25).

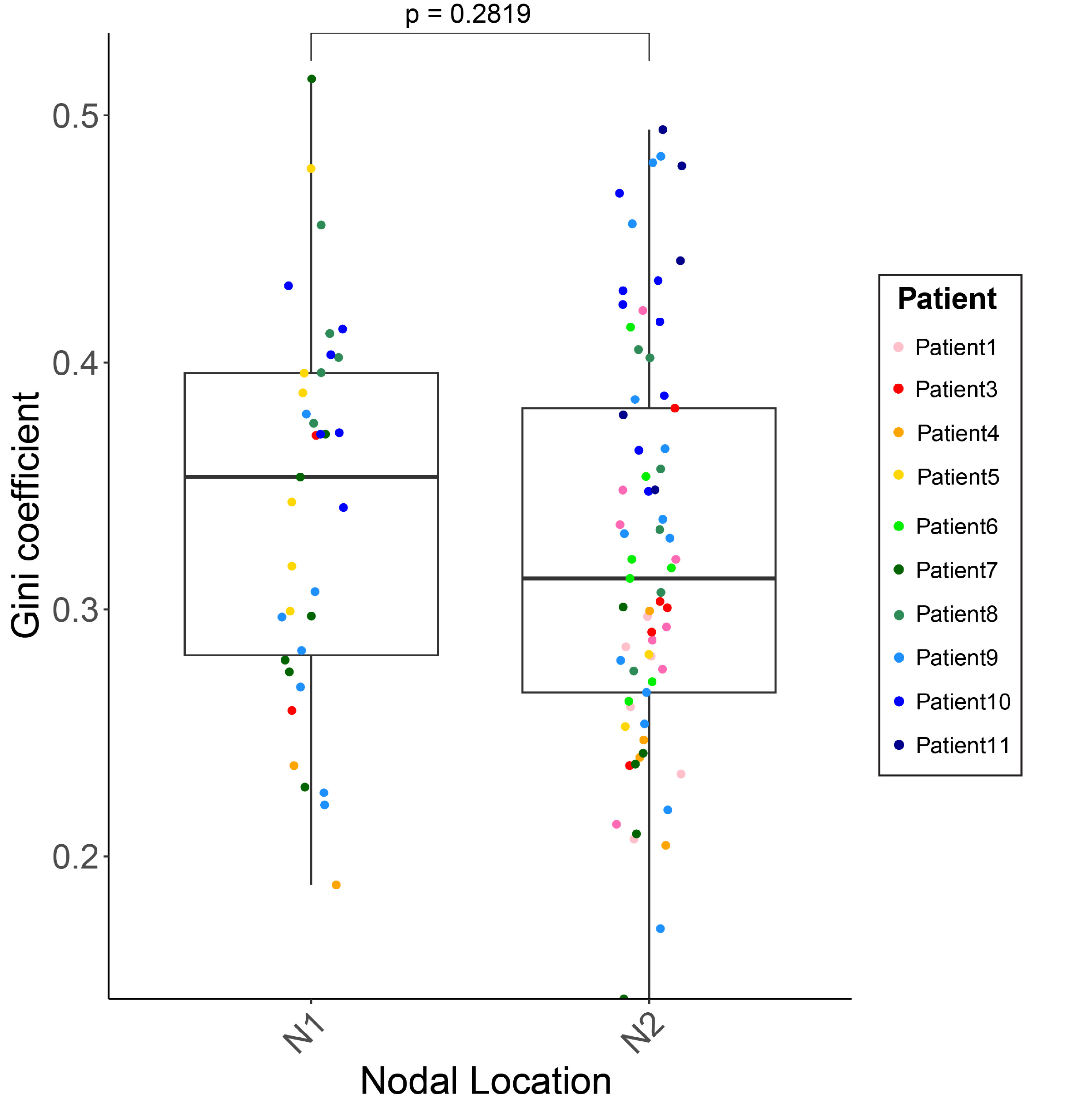

**Supplementary Figure 5. Assessing variability of cell type compositions from the IMC data across nodal regions.** To measure the variability in cell type composition derived by scRNA-seq/CITE-seq, we calculated the Gini coefficient for each sample based on the proportions of the eight identified broad cell types (T/NK, B, plasma, myeloid, pDC, neutrophil, mast cell, and epithelial). A lower Gini coefficient indicates less variability of the cell types within the sample. Each dot represents a lymph node and is color-coded by the originating patient. No statistically significant differences were observed in the Gini coefficient between broad cell types across the two nodal locations. The P-value was computed using a t-test between nodal regions.

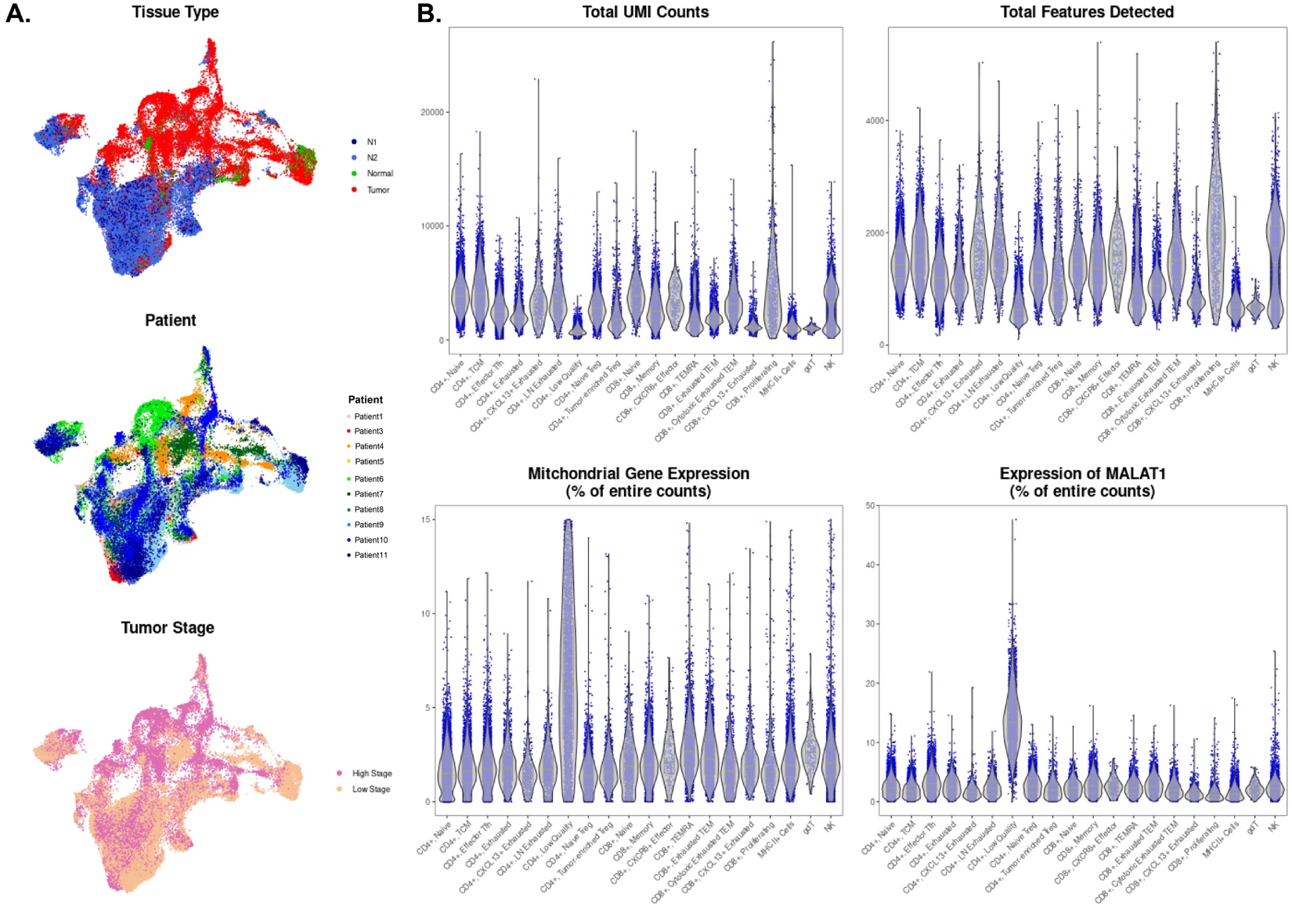

**Supplementary Figure 6. Quality control metrics for 44,243 T lymphocyte and NK cells.  A)** UMAP projections generated on the T lymphocyte and NK cell populations are color-coded by the originating tissue type, originating patient, and the tumor stage of the originating patient. **B)** The total UMI counts, total features detected, the mitochondrial gene expression percentage and the MALAT1 expression percentage are shown for each T/NK cell population.

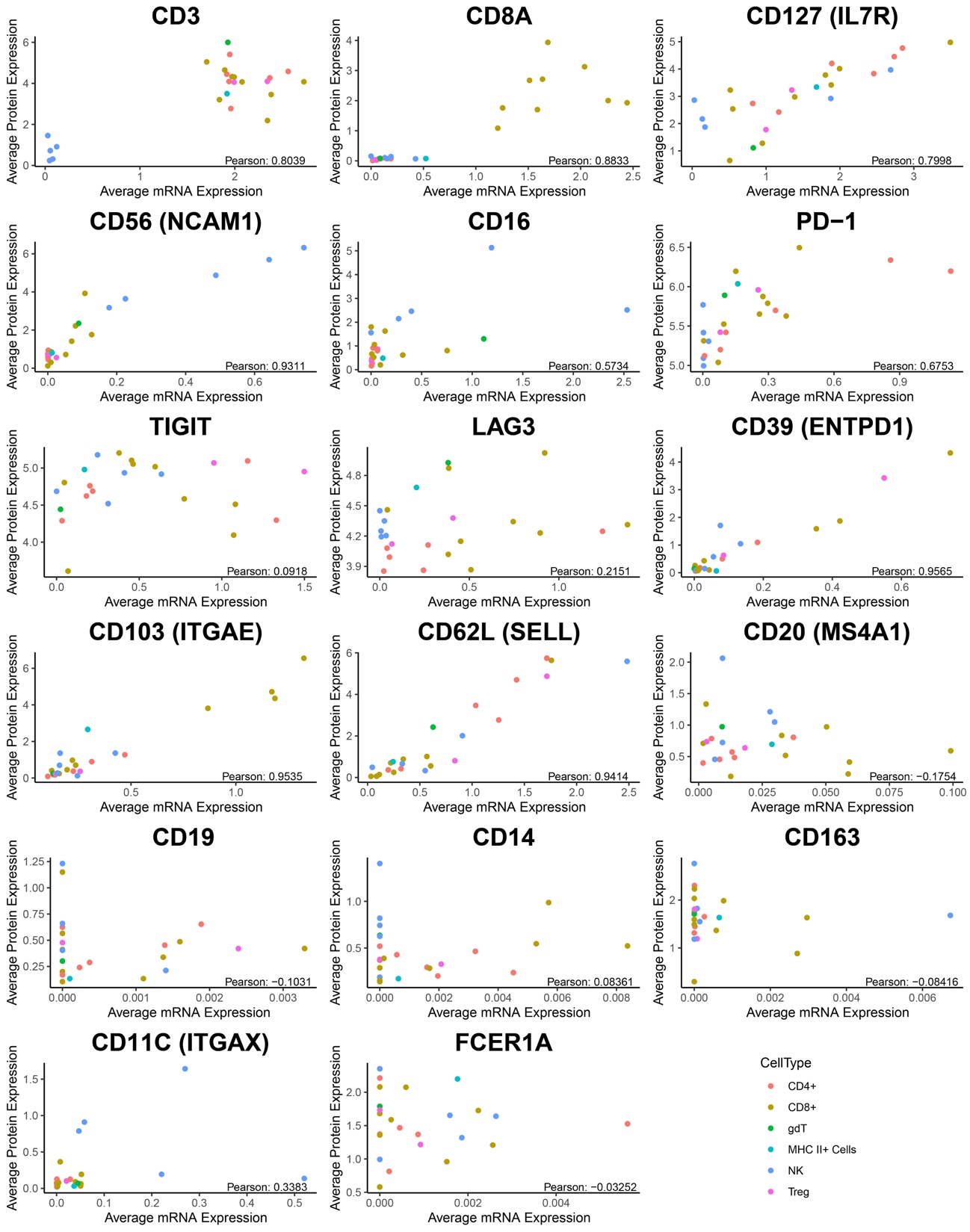

**Supplementary Figure 7. Comparison of mRNA and corresponding protein expression across T lymphocytes and NK cells in scRNA-seq/CITE-seq data.** Scatterplots display the average mRNA (x-axis) and corresponding protein (y-axis) expression levels of selected markers across 21 T/NK subtypes. Each dot represents an identified cell subtype and is color-coded by the broad cell type: CD4+, Treg, CD8+, NK, or Other. Other includes the γδ population and an MHC-II+ CD4+/CD8+ population. The calculated Pearson Correlation Coefficient is shown on the bottom right. While CD4, CD8, and markers associated with T lymphocyte activation (CD39, CD103) and homing (CD62L, IL7R) display high correlation, markers associated with T lymphocyte exhaustion exhibited low correlation (< 0.25).

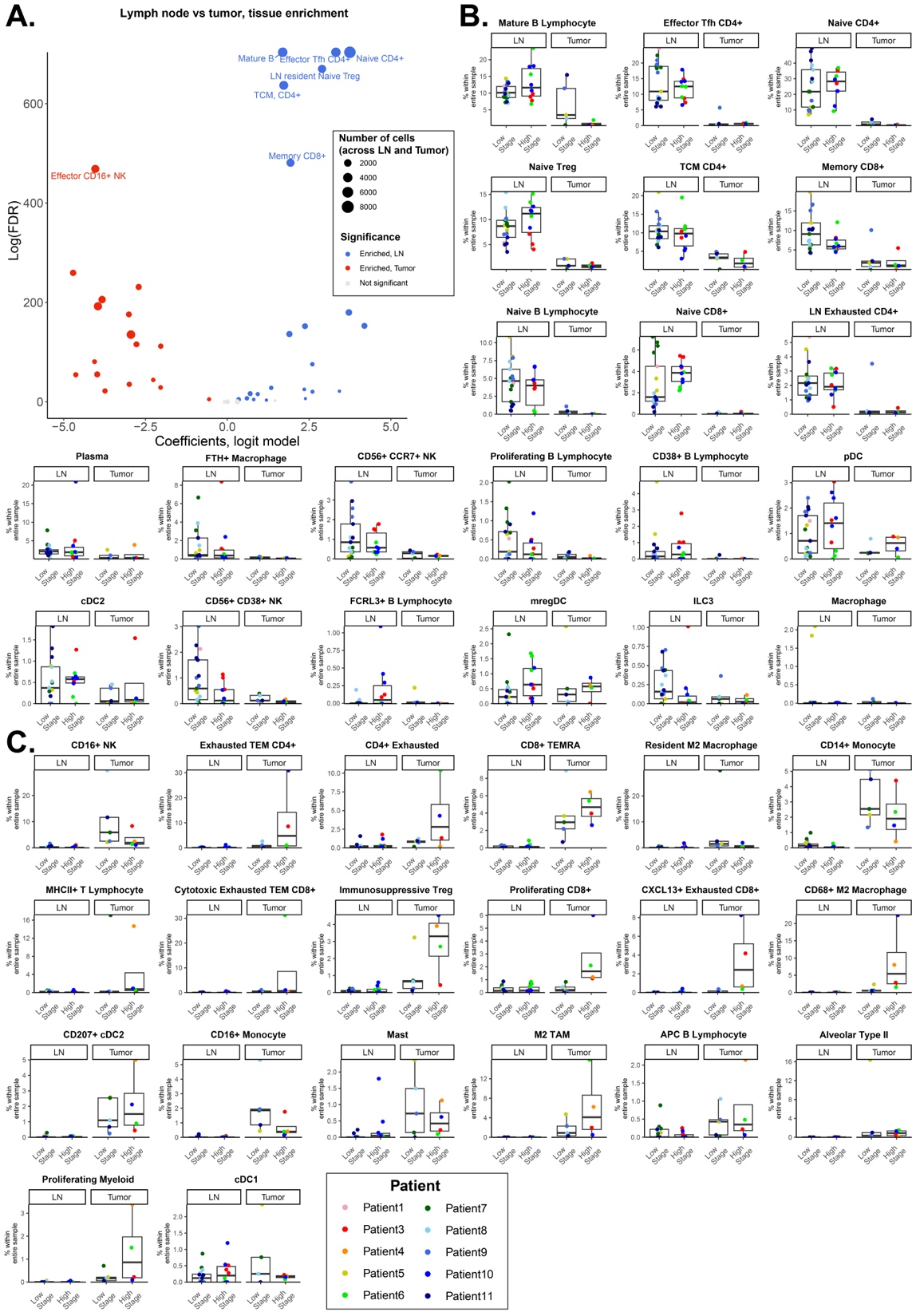

**Supplementary Figure 8. Association of cell populations from scRNA-seq/CITE-seq with tissue type.**  **A)** A logistic regression model was applied to determine which cell populations were enriched in lymph node of tumor tissue while controlling for originating patient and tumor stage. 21 cell populations were enriched in the lymph nodes while 20 cell populations were enriched in the tumor tissue (FDR < 0.05). Each dot represents a cell population. Dot sizes represent the total number of cells in the cell population across all lymph nodes. **B)** The proportions of cell populations that were enriched in lymph nodes are shown as a function of stage and tissue type. **C)** The proportions of cell populations that were enriched in tumor tissue are shown as a function of stage and tissue type. Each data point represents a sample. The color of each point indicates the patient from which the sample was derived.

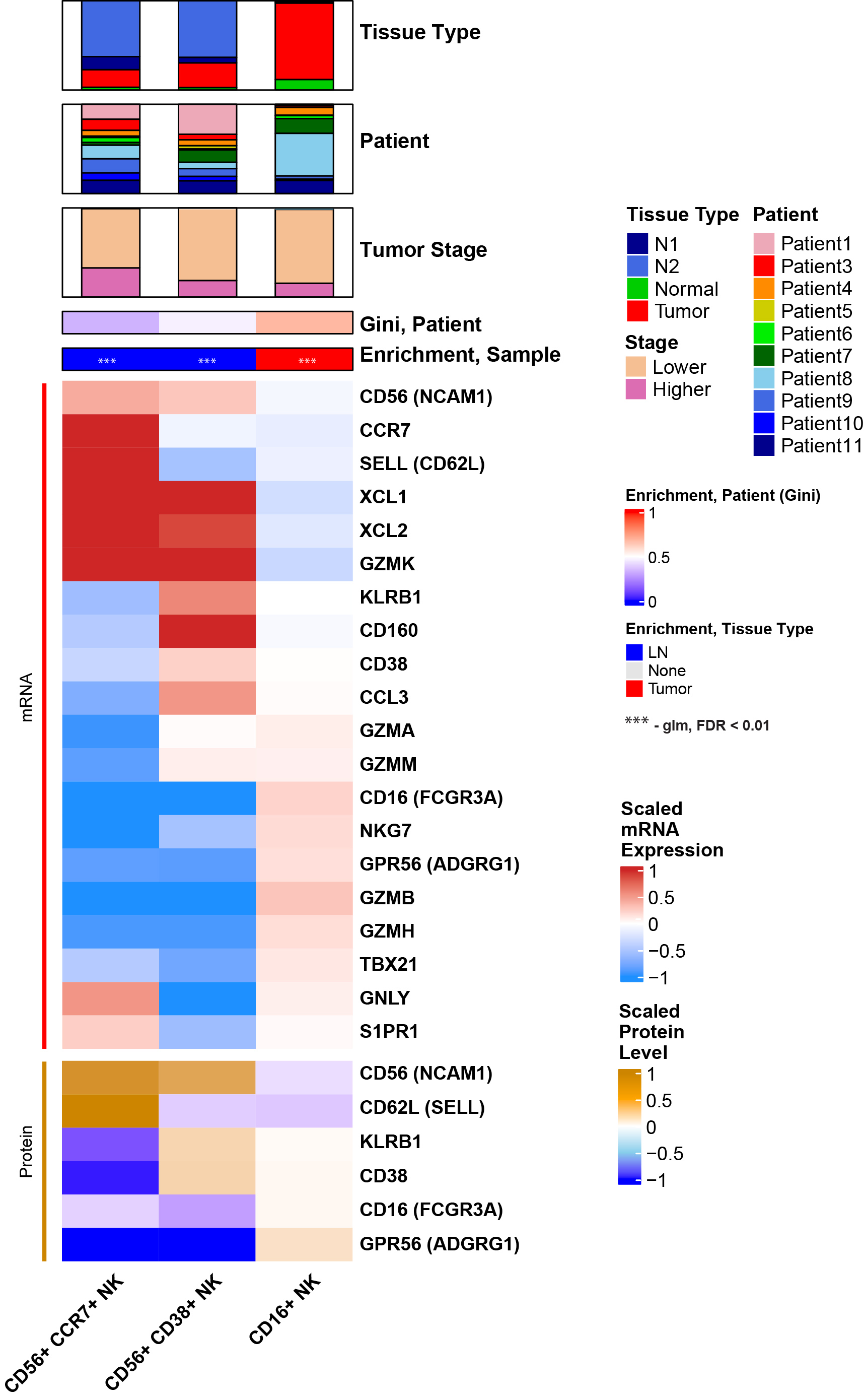

**Supplementary Figure 9. Multimodal characterization of NK cell subpopulations.** Expression of selected mRNA and protein markers are shown for the 3 NK cell subpopulations. All markers were differentially expressed (FDR < 0.05, Log2FoldChange > 0.25) in their respective subpopulation when compared against other T lymphocyte and NK cell populations. Expression is scaled across all NK cells and averaged per NK cell cluster.

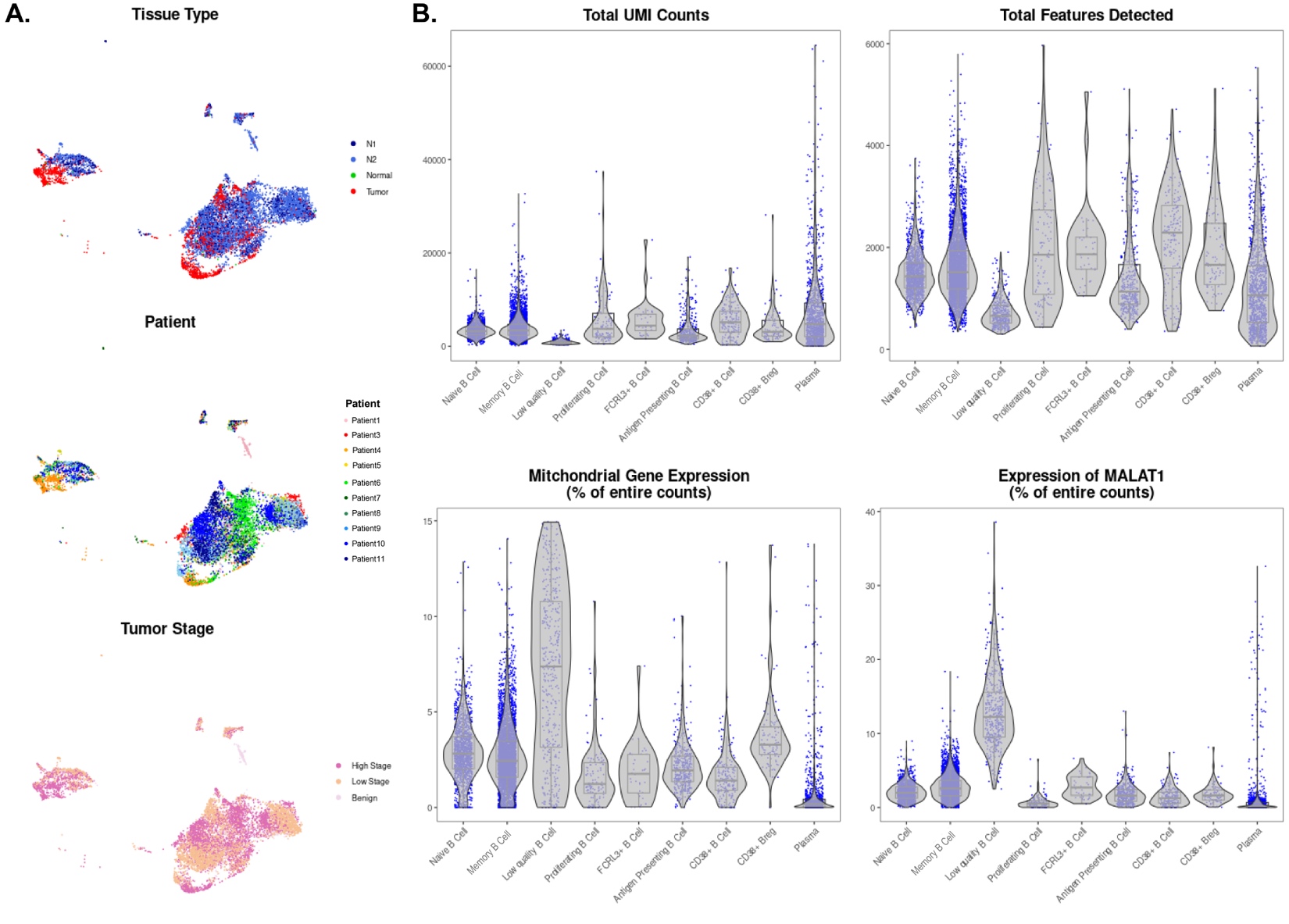

**Supplementary Figure 10. Quality control metrics for 9,099 B lymphocytes and plasma cells.** **A)** UMAP projections generated on the B lymphocyte and plasma cell populations are color-coded by the originating tissue type, originating patient, and the tumor stage of the originating patient. **B)** The total UMI counts, total features detected, the mitochondrial gene expression percentage, and the MALAT1 expression percentage are shown for each cell population.

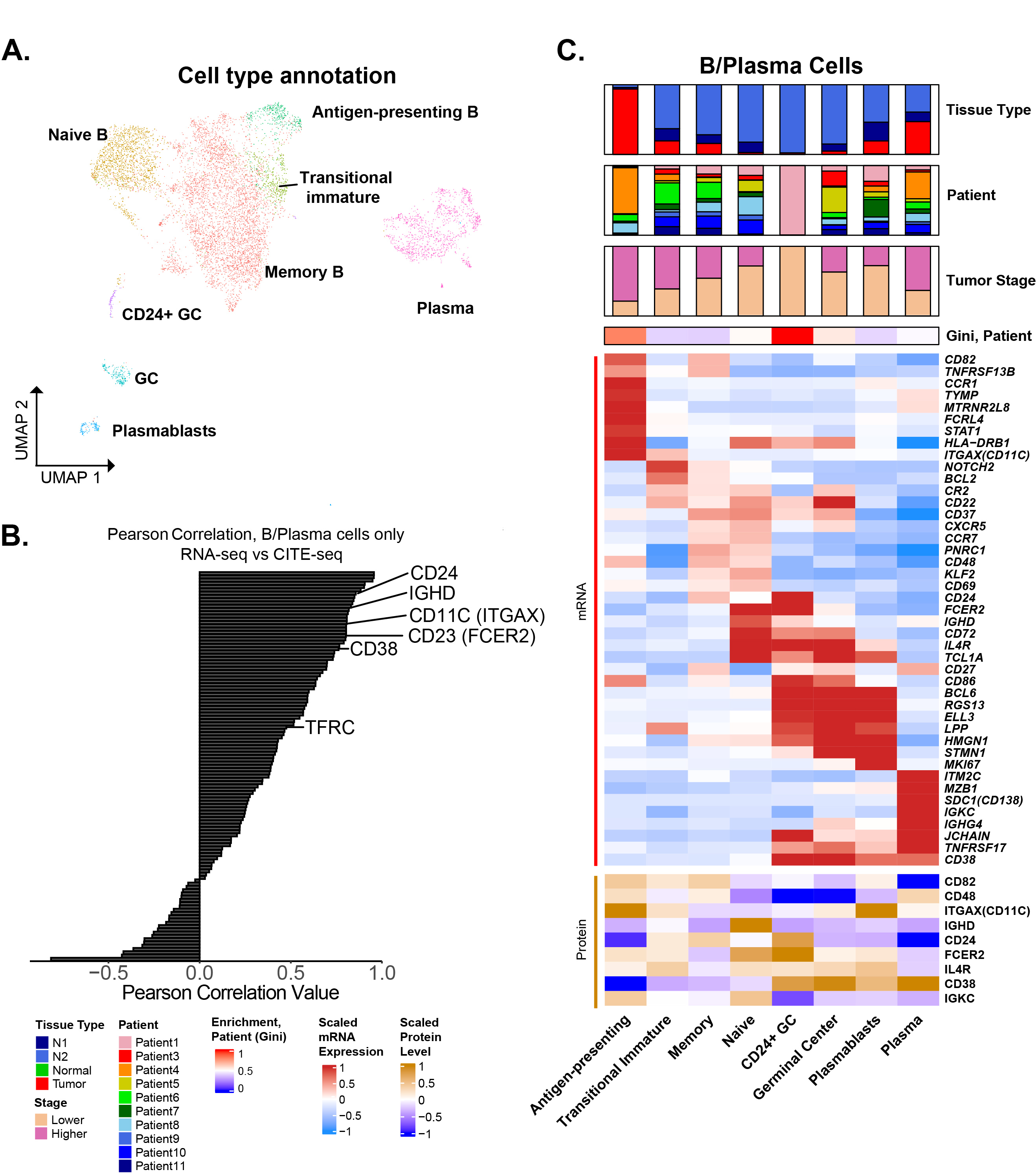

**Supplementary Figure 11. Multimodal characterization of B-cell and plasma cell subpopulations. A)** RNA expression-based UMAP projection of B cells and plasma cells annotated by subpopulation. **B)** The Pearson correlation coefficient was calculated across 122 markers in the CITE-seq panel within the B cell/plasma cell populations. Selected markers associated with B-cell/plasma cell populations are shown. **C)** Expression of RNA and protein markers associated with each B-cell/plasma cell subpopulation is shown. Expression for each marker was scaled across all B/plasma cells and then averaged per subpopulation.

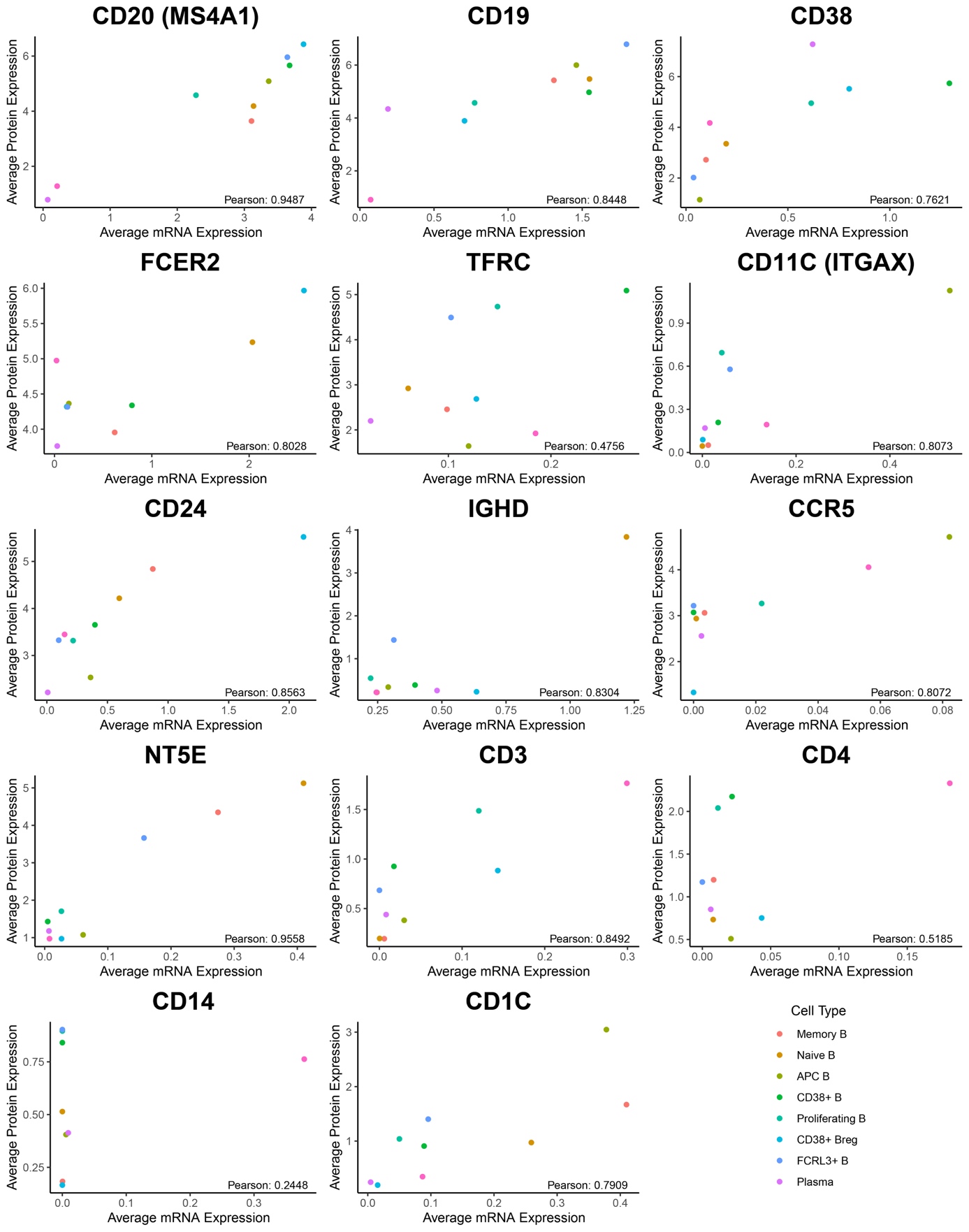

**Supplementary Figure 12. Comparison of mRNA and corresponding protein expression across B lymphocytes in scRNA-seq/CITE-seq data.** Scatterplots display the average mRNA (x-axis) and corresponding protein (y-axis) expression levels of selected markers across a total of 8 B/plasma cell subtypes. Each dot represents an identified cell subtype and is color-coded a B lymphocyte or plasma cell. The calculated Pearson Correlation Coefficient is shown on the bottom right. Markers for B lymphocytes and plasma cells, such as CD19, CD20, and CD38, have a high correlation between RNA and protein expression.

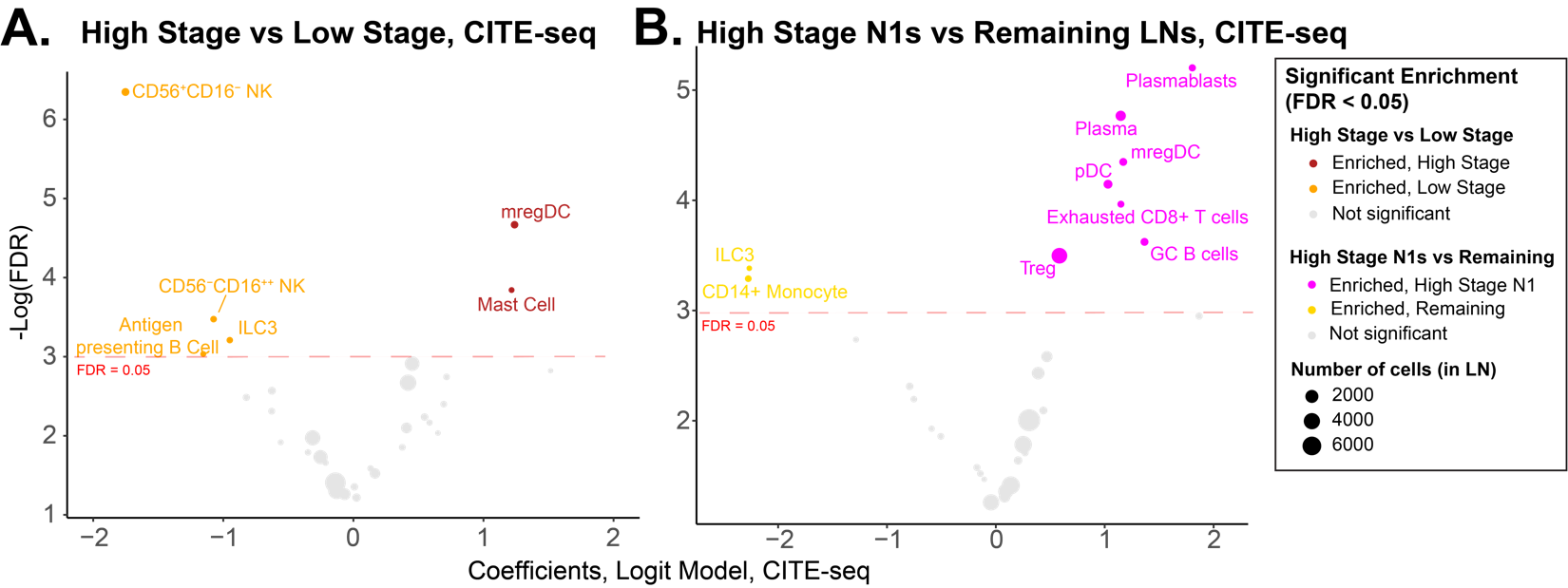

**Supplementary Figure 13. Association of cell populations from scRNA-seq/CITE-seq with stage and nodal region*.***  **A)** A logistic regression model was applied to identify cell populations from the scRNA-seq/CITE-seq data that were enriched in lower or higher-stage patients while controlling for originating patient and nodal region. 4 cell populations were enriched in LNs of lower-stage patients, and 2 cell populations were enriched in the those of high-stage patients (FDR < 0.05). Each dot represents a cell population. Dot sizes represent the total number of cells in the cell population across all lymph nodes. **B)** A similar logistic regression model was applied to compare N1 regions of higher-stage patients against all other lymph nodes. Multiple immune populations with known immunosuppressive function, including Tregs, exhausted CD8^+^ T cells and mregDCs, were enriched in the N1 LNs of higher-stage patients (FDR < 0.05).

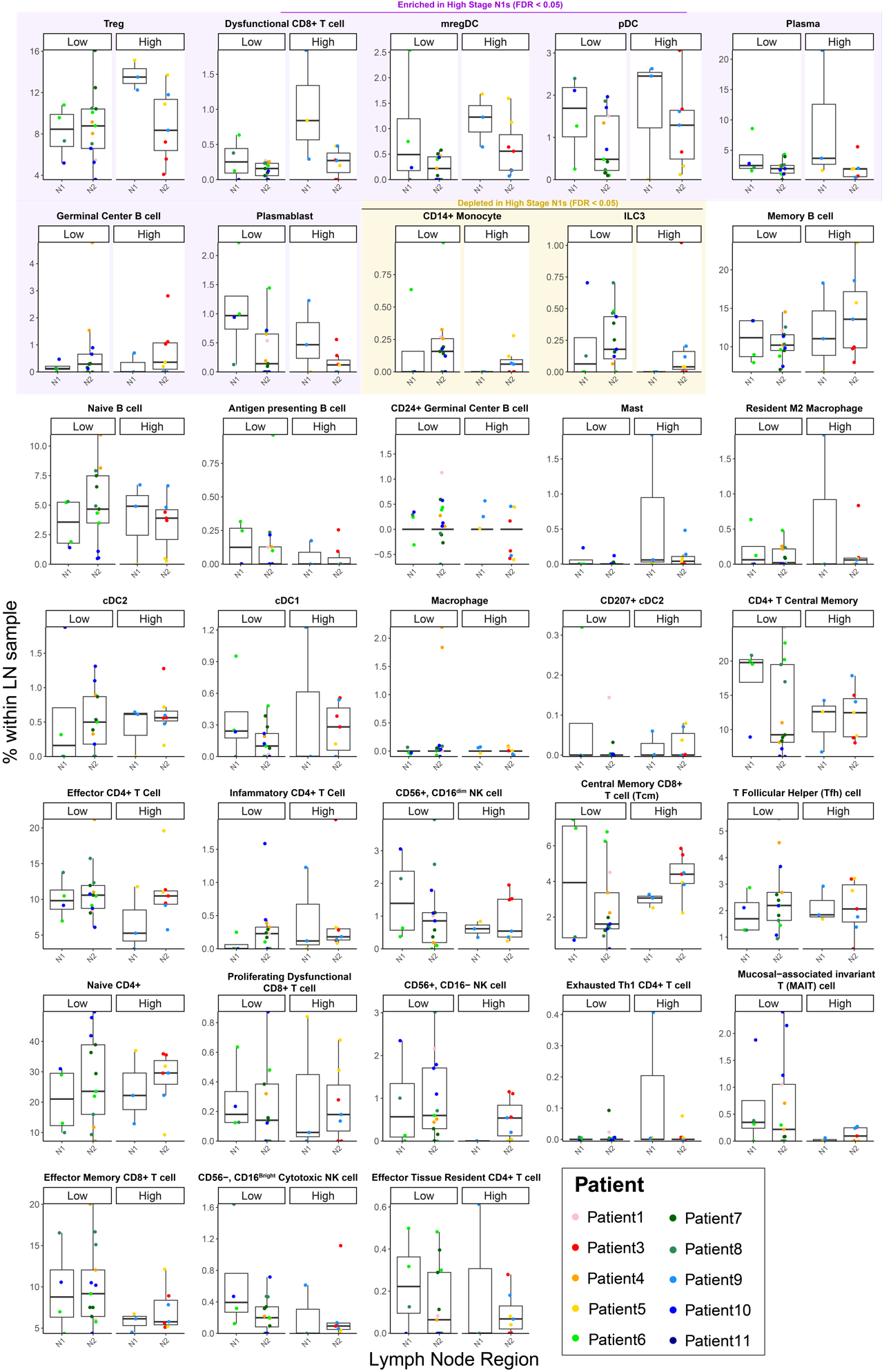

**Supplementary Figure 14. Association of cell populations from scRNA-seq/CITE-seq with N1 LNs in higher-stage patients**. The proportions of cell populations that were enriched or depleted in N1 LNs of higher-stage patients are shown as a function of stage and nodal region. Each data point represents a lymph node, with colors of the dot indicating the respective patient the sample was collected from. Cell populations significantly enriched or depleted in N1 lymph nodes of high-stage patients are highlighted in purple and yellow, respectively (FDR < 0.05).

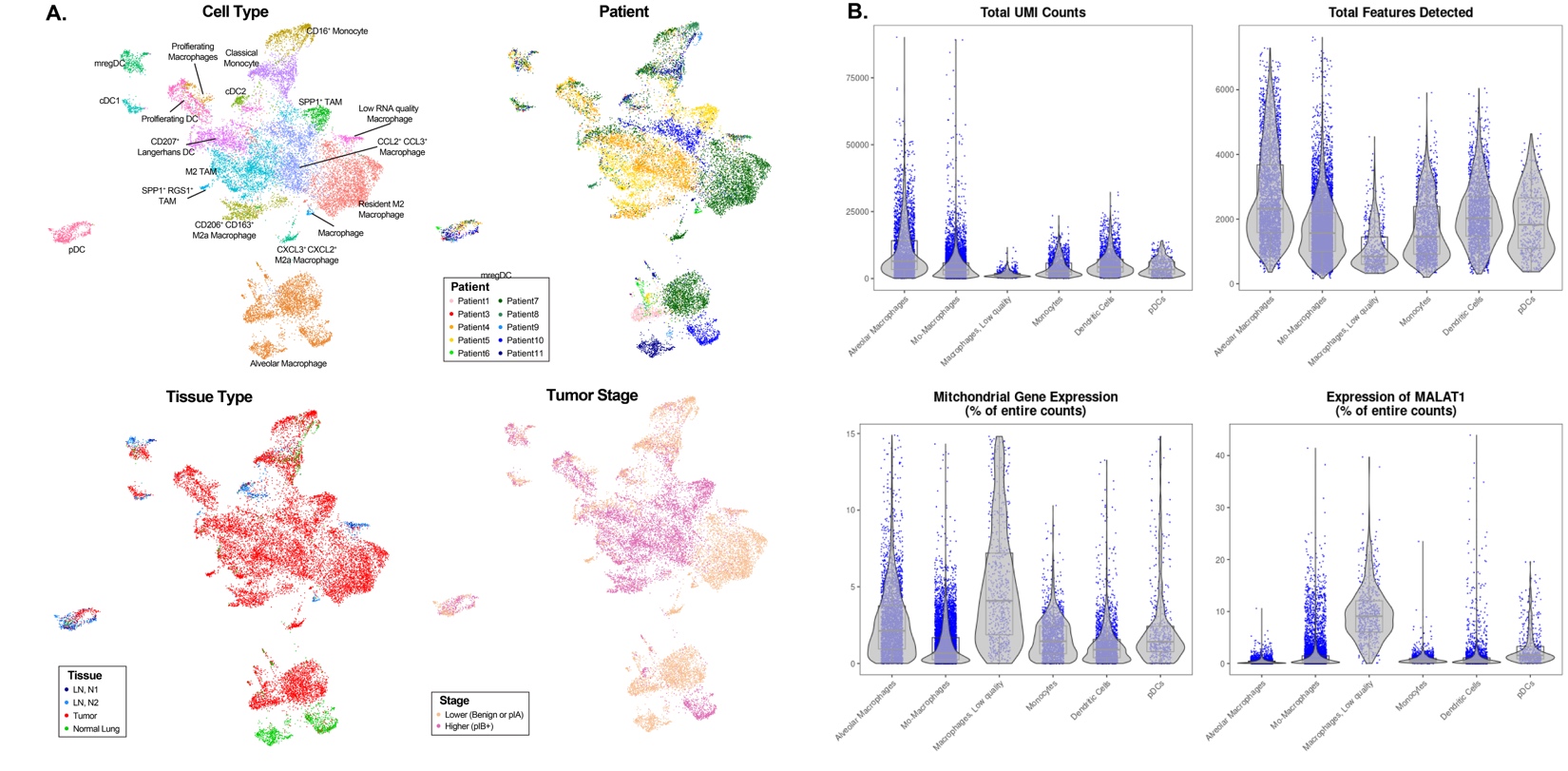

**Supplementary Figure 15. Quality control metrics for 17,392 myeloid cells.**   **A)** UMAP projections generated on the myeloid cell populations are color-coded by the originating tissue type, originating patient, and the tumor stage of the originating patient. **B)** The total UMI counts, total features detected, the mitochondrial gene expression percentage and the MALAT1 expression percentage are shown for each myeloid cell population.

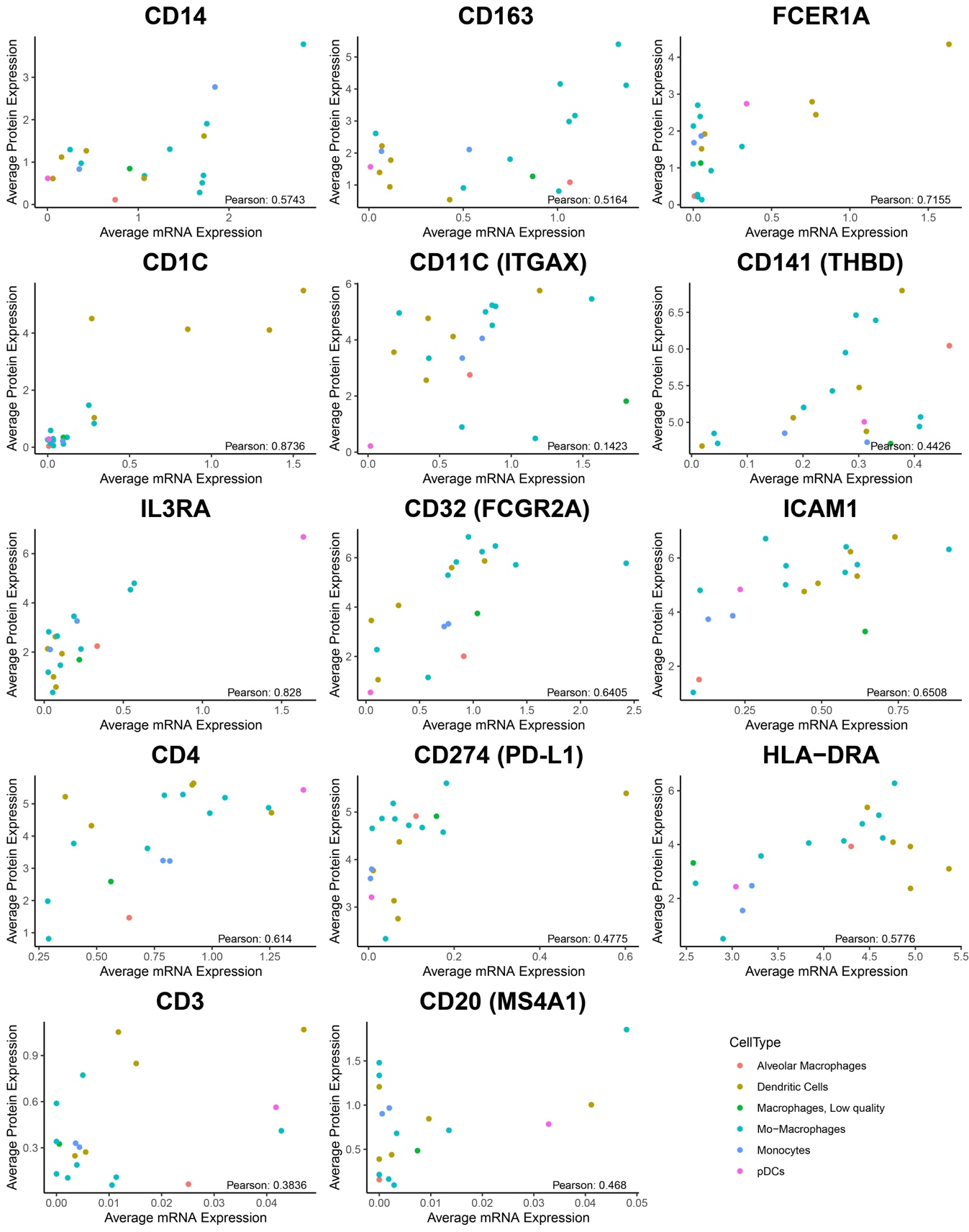

**Supplementary Figure 16. Comparison of mRNA and corresponding protein expression across myeloid cells.** Scatterplots display the average mRNA (x-axis) and corresponding protein (y-axis) expression levels of selected markers across 19 myeloid cell subtypes. Each dot represents an identified cell subtype and is color-coded per myeloid cell type. The calculated Pearson Correlation Coefficient is shown on the bottom right. While some dendritic cell markers such as CD1C and FCER1A show a high correlation between modalities, other markers such as CD11C have lower correlations across modalities.

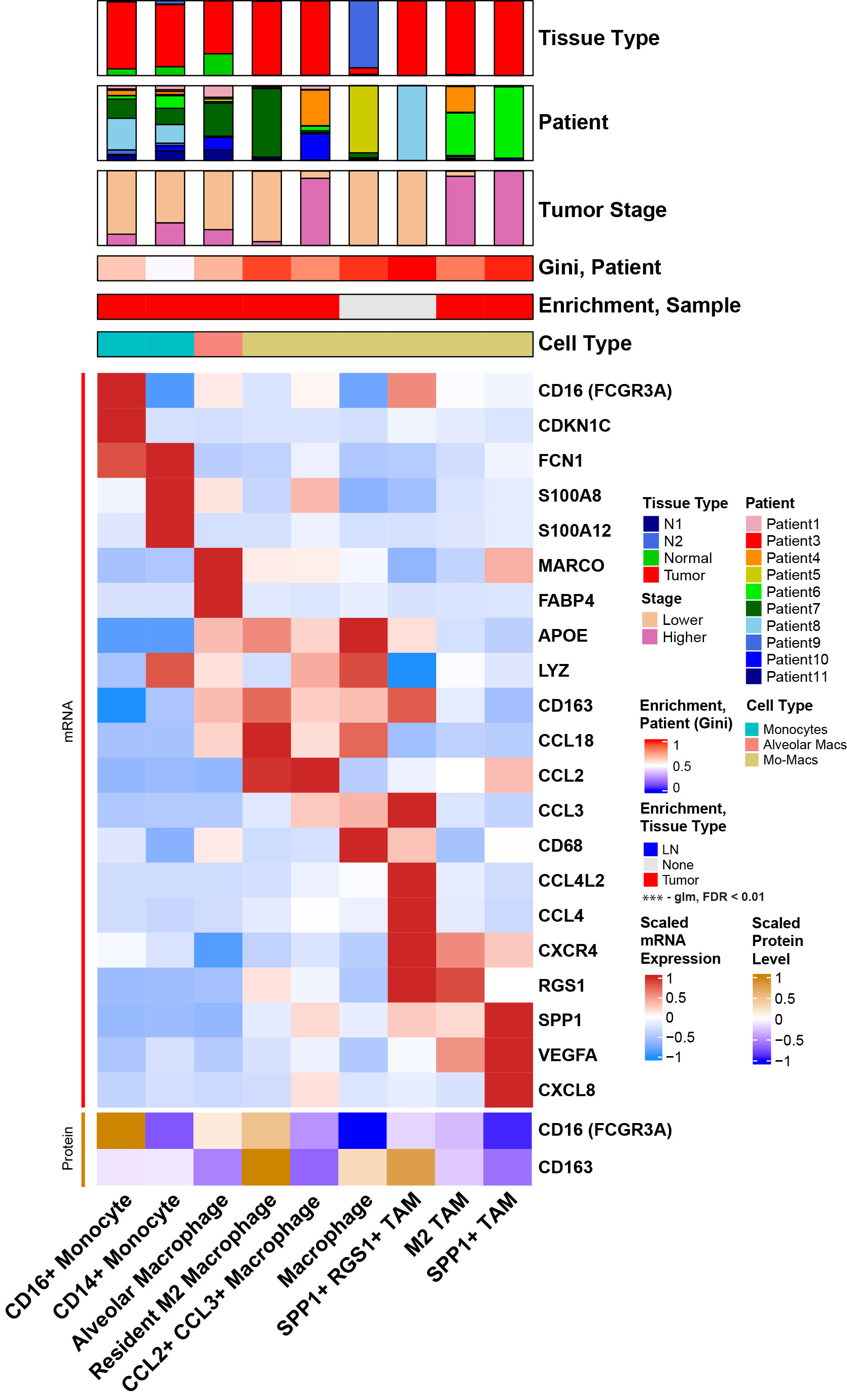

**Supplementary Figure 17. Multimodal characterization of macrophage and monocyte subpopulations.** Expressions of selected RNA and protein markers are shown for macrophage and monocyte subpopulations. All markers were differentially expressed for their respective subpopulation when compared against other myeloid cell populations (FDR < 0.05, Log2FoldChange > 0.25). Expression for each marker was scaled across all myeloid cells and the averaged per cell subpopulation.

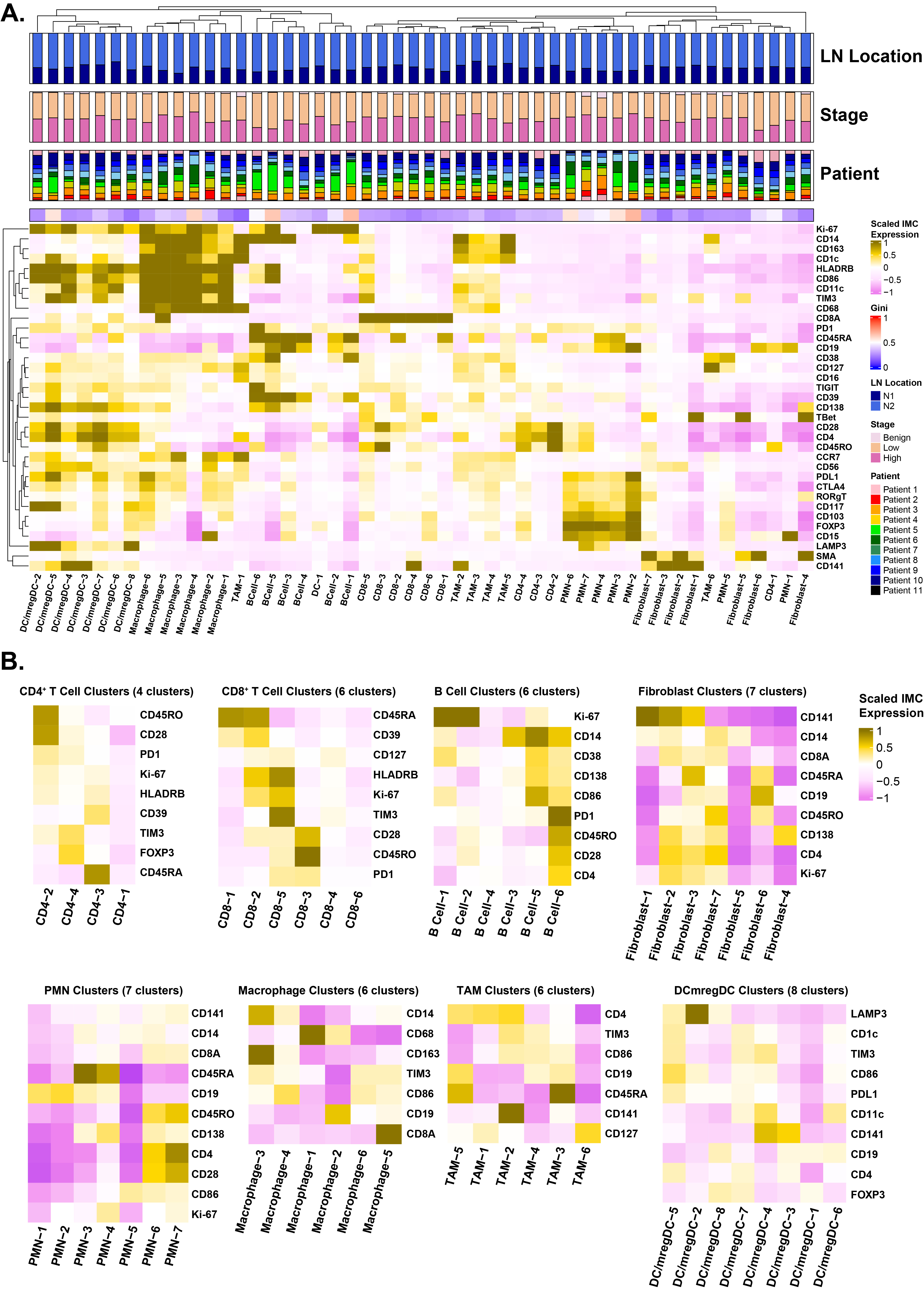

**Supplementary Figure 18. Immune cell subpopulations were identified through subclustering of IMC-defined cell types.  A)** Each of the 8 IMC-based cell types, including Macrophage, TAM, DC, CD4^+^ T Cell, CD8^+^ T Cell, B Cell, PMN, Fibroblast, were further subclustered using Celda. Antibody expression for all markers was Z-score normalized across all cells and averaged for each of the 50 subclusters. **B)** Antibody expression for selected markers was Z-score normalized for the cells within each major cell type and averaged for each of the subclusters within a major cell type. The expression of these markers was used to define the cell subpopulations and/or potentially neighboring cell interactions.

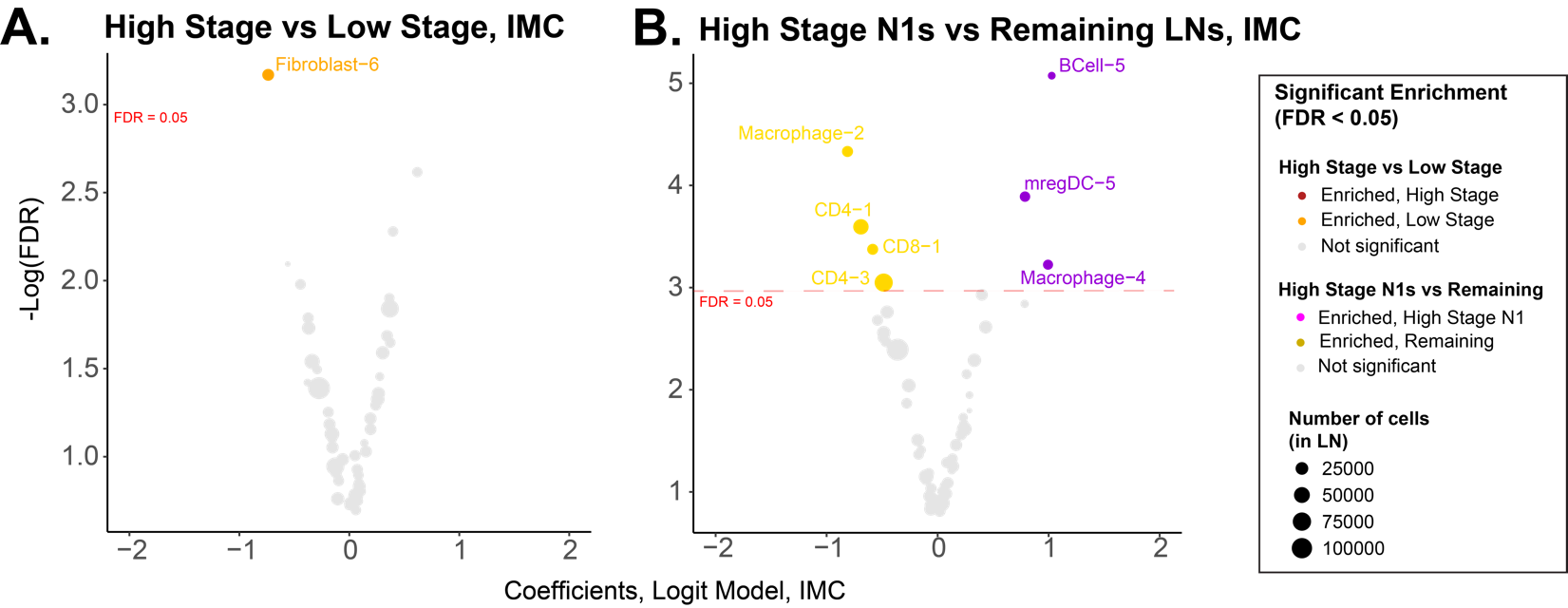

**Supplementary Figure 19. Association of IMC-based immune cell cluster abundance to higher stage phenotypes*.*** Logistic regression models controlling for patient and tumor stage was applied to identify IMC based cell to identify clusters enriched in **A)** patients with higher tumor stage compared to lower tumor stage or **B)** enriched in the N1 LNs of high-stage patients compared to all other LNs (FDR < 0.05).

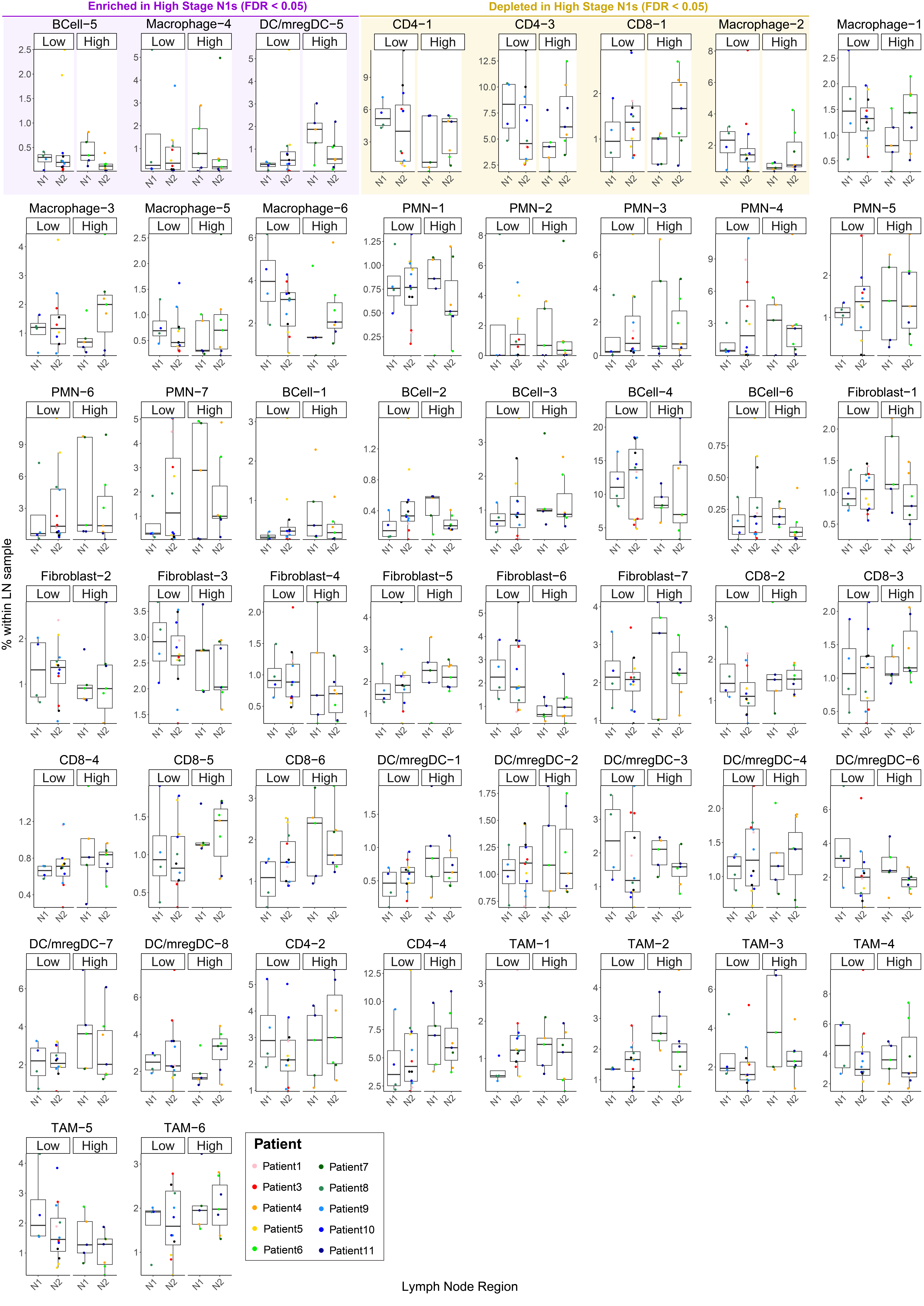

**Supplementary Figure 20. Proportions of IMC cell clusters across nodal regions and stage groups.**  The proportions of IMC-based cell clusters are depicted per lymph node across nodal regions and stage groups. Each data point represents the proportion of a cell cluster in a single lymph node colored by the respective patient from which the sample was collected. Clusters enriched or depleted in higher-stage N1 lymph nodes are highlighted in purple (enriched in higher-stage N1 LNs) or yellow (depleted in higher-stage N1 LNs). Abbreviations: High: Higher-stage, Low: Lower-stage.

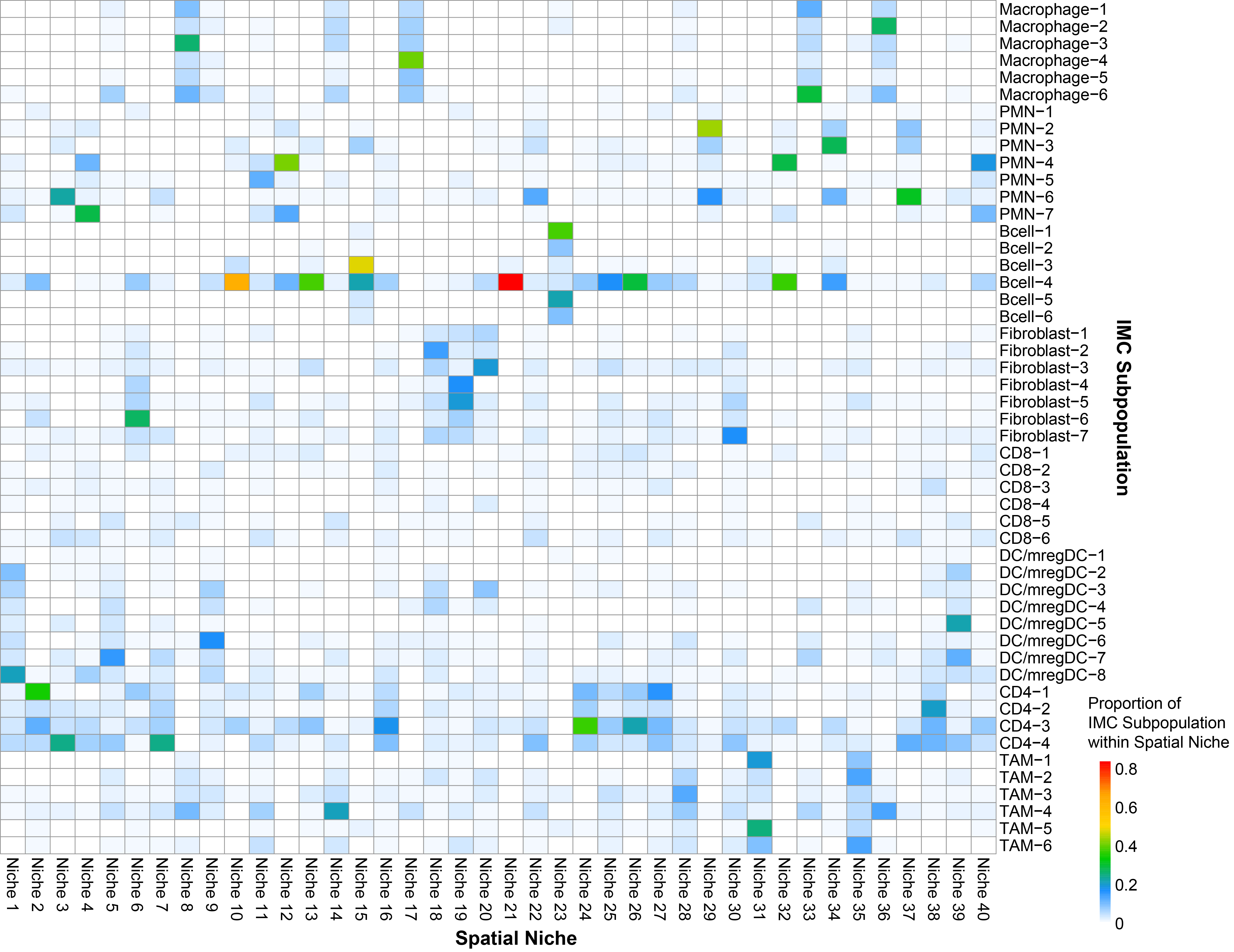

**Supplementary Figure 21. Proportion of IMC clusters within Spatial Niches.**  For each cell within the IMC dataset, the number of neighboring cells from each cell cluster within a 20µm radius was quantified and used to define spatial niches. 40 niches were generated in total, each representing a unique spatial neighborhood of immune and stromal cell subpopulations. The proportion of each IMC cell cluster is shown for each spatial niche.

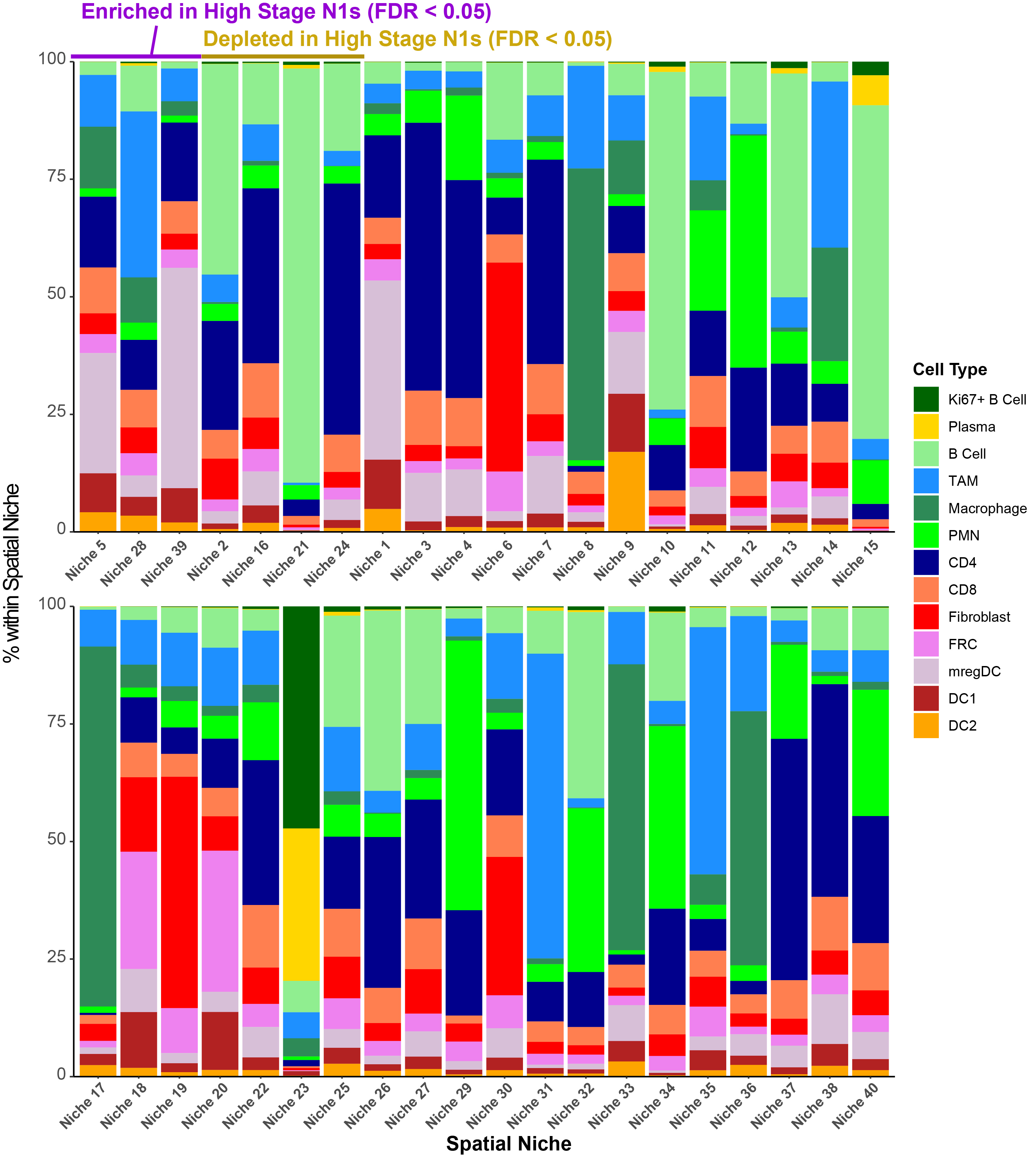

**Supplementary Figure 22. Composition of cell types within spatial niches.** Spatial niches are comprised of a mixture of immune and stromal cell populations co-occurring across the lymph node samples. Proportions of the major cell types within each niche are shown. Niches enriched (purple) or depleted (yellow) in N1 LNs from higher-stage patients are labeled accordingly (FDR < 0.05).

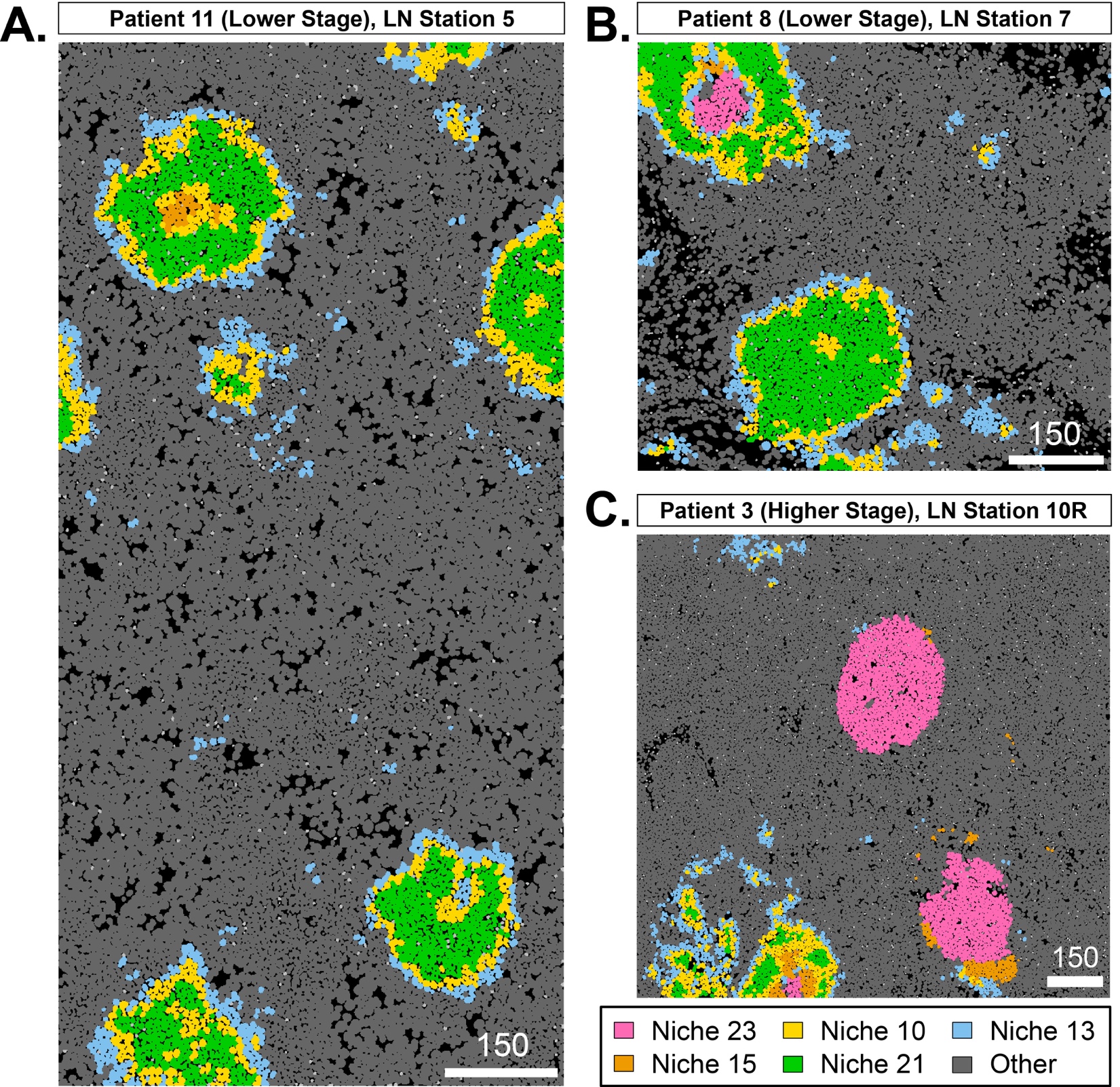

**Supplementary Figure 23. Characterization of B cell follicles through spatial arrangement of niches.** **A**) The spatial patterns of spatial niches 23 (representing the germinal center of B cell follicles) and niches 15, 10, 21, and 13 (representing the mantle zone) were used to identify B cell follicles. In lower-stage lymph nodes (LNs), dormant B cell follicles lacking a germinal center were preferentially observed. **B)** Amongst B cell follicles containing a germinal center, those present in lower-stage lymph nodes had a higher likelihood of being fully encapsulated by the mantle zone. **C)** Higher-stage lymph nodes were enriched for B cell follicles which partially or fully lacked a mantle zone encapsulating the germinal center.

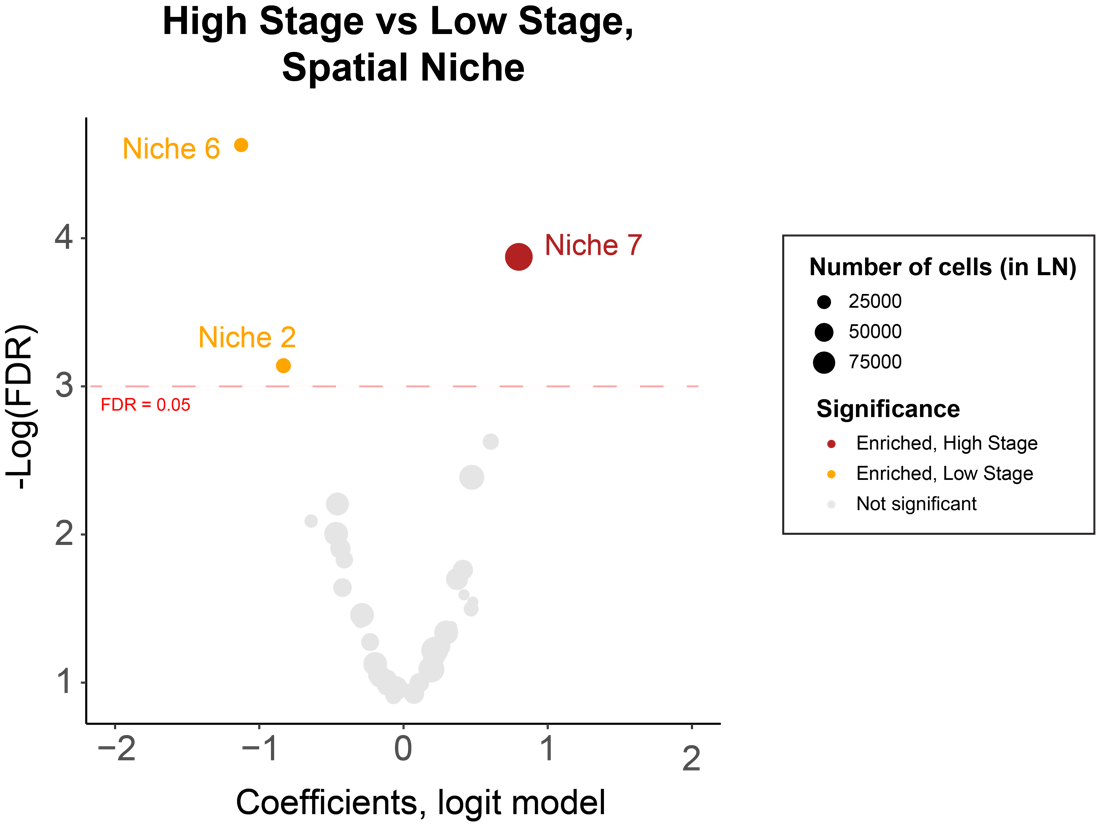

**Supplementary Figure 24. Enrichment of IMC spatial niches by tumor stage*.*** A logistic regression model controlling for originating patient and nodal region was applied to identify spatial niches enriched in LNs of either lower or higher-stage patients. (FDR < 0.05). Each dot represents a spatial niche. Dot sizes represent the total number of cells in the niche across all lymph nodes.

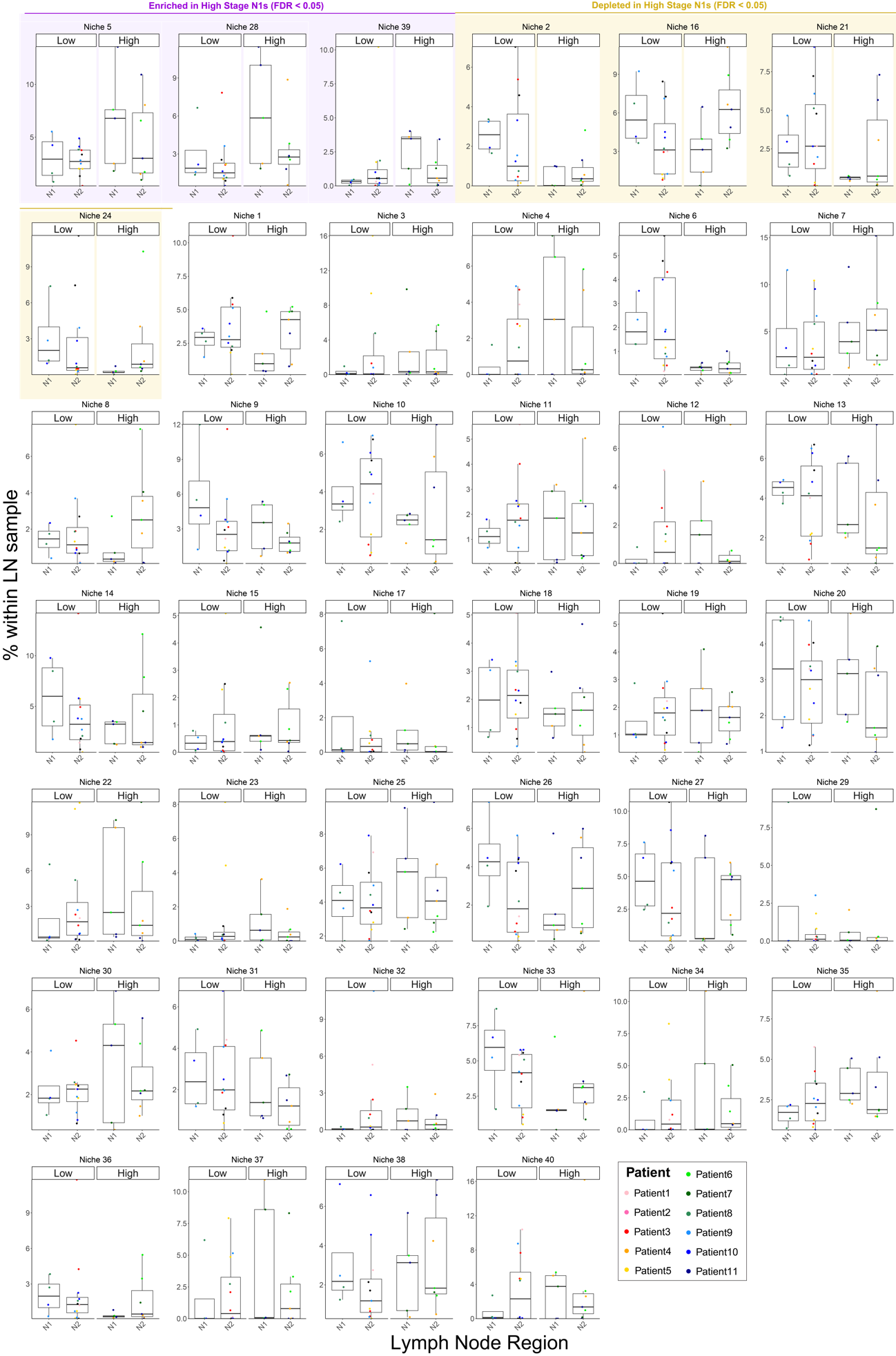

**Supplementary Figure 25. Determining nodal enrichment of imaging-based spatial niches*.***  The proportions of spatial niches per sample are depicted. Each data point represents a lymph node, with colors indicating the respective patient the sample was collected from. Niches significantly enriched or depleted in N1 lymph nodes of high-stage patients are highlighted in purple and yellow, respectively (FDR < 0.05).

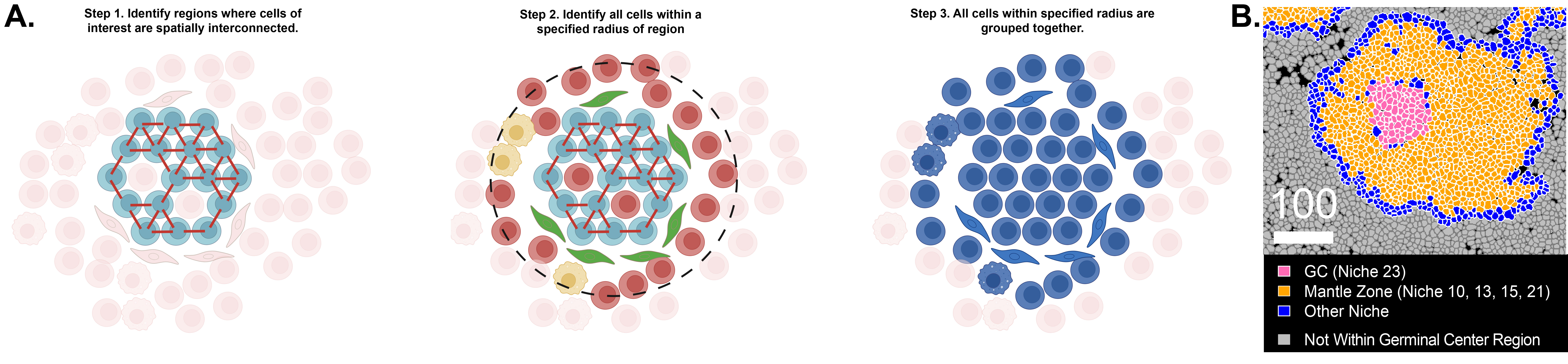
**Supplementary Figure 26. Computationally defining B cell follicular regions*.***  **A**) A computational approach was taken to define germinal center and B cell follicular regions. **B)** Single-cell masks for cells that were identified as part of the germinal center region are colored accordingly. All other cells are shaded gray. *Lymph node: Patient 10, LN Station 10R, ROI 1*

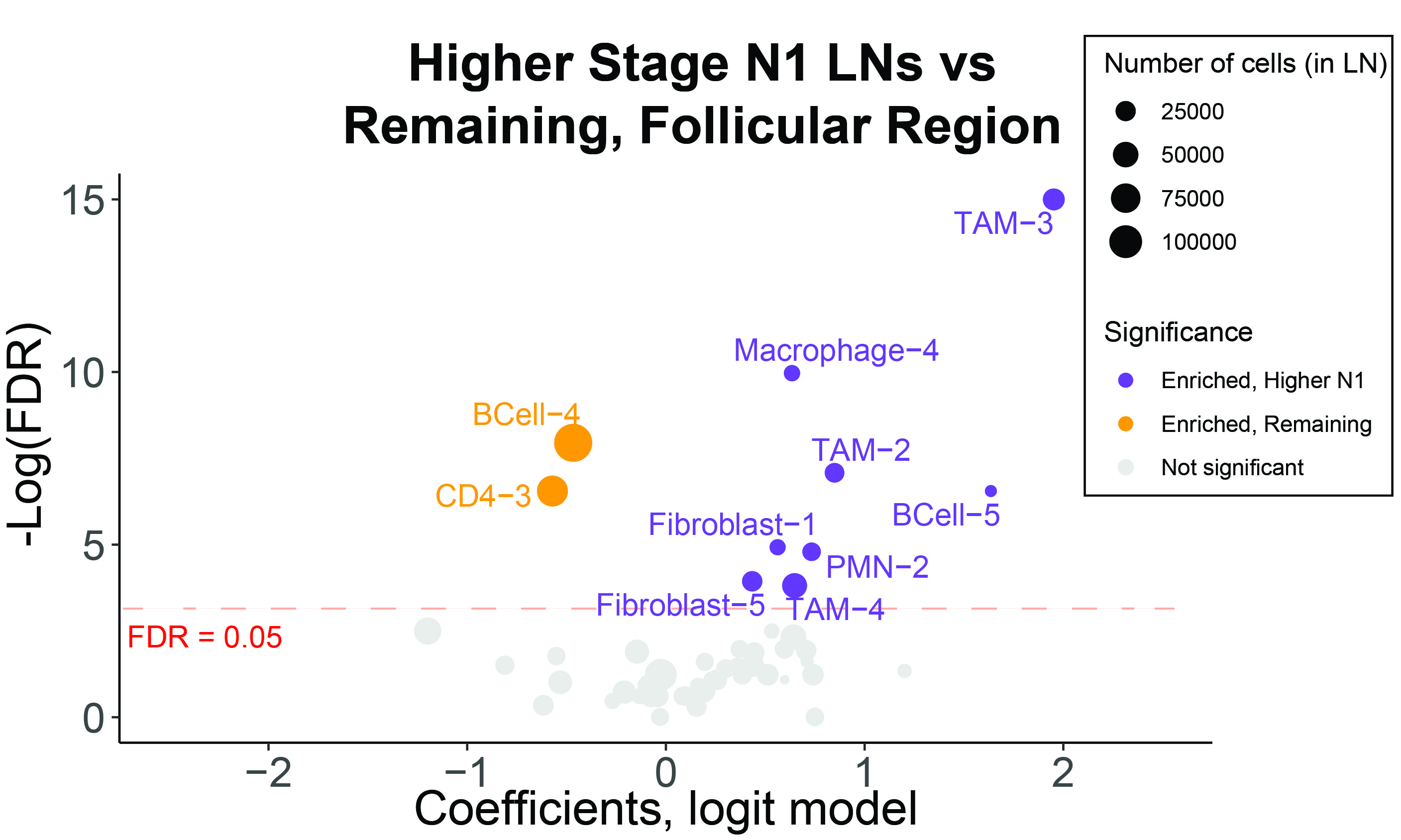
**Supplementary Figure 27. Comparison of immune cell populations with B cell follicular regions by nodal region.** A logistic regression model controlling for originating patient per cell and tumor stage was applied to identify IMC based cell clusters that were enriched or depleted in computationally defined follicle regions of higher-stage N1 lymph nodes.
